# The second wave of SARS-CoV-2 infections and COVID-19 deaths in Germany – driven by values, social status and migration background? A county-scale explainable machine learning approach

**DOI:** 10.1101/2021.04.14.21255474

**Authors:** Gabriele Doblhammer, Constantin Reinke, Daniel Kreft

## Abstract

There is a general consensus that SARS-CoV-2 infections and COVID-19 deaths have hit lower social groups the hardest, however, for Germany individual level information on socioeconomic characteristics of infections and deaths does not exist. The aim of this study was to identify the key features explaining SARS-CoV-2 infections and COVID-19 deaths during the upswing of the second wave in Germany.

We considered information on COVID-19 diagnoses and deaths from 1. October to 15. December 2020 on the county-level, differentiating five two-week time periods. We used 155 indicators to characterize counties in nine geographic, social, demographic, and health domains. For each period, we calculated directly age-standardized COVID-19 incidence and death rates on the county level. We trained gradient boosting models to predict the incidence and death rates with the 155 characteristics of the counties for each period. To explore the importance and the direction of the correlation of the regional indicators we used the SHAP procedure. We categorized the top 20 associations identified by the Shapley values into twelve categories depicting the correlation between the feature and the outcome.

We found that counties with low SES were important drivers in the second wave, as were those with high international migration and a high proportion of foreigners and a large nursing home population. During the period of intense exponential increase in infections, the proportion of the population that voted for the Alternative for Germany (AfD) party in the last federal election was among the top characteristics correlated with high incidence and death rates.

We concluded that risky working conditions with reduced opportunities for social distancing and a high chronic disease burden put populations in low-SES counties at higher risk of SARS-CoV-2 infections and COVID-19 deaths. In addition, noncompliance with Corona measures and spill-over effects from neighbouring counties increased the spread of the virus. To further substantiate this finding, we urgently need more data at the individual level.

## INTRODUCTION

The second wave of SARS-CoV-2 infections has hit Germany, starting in October, picking up exponential speed in November and remaining at a high plateau during much of December despite lockdown measures from the beginning of December onwards. Information about gender, age, and place of residence was still the only available information about SARS-CoV-2 infections and COVID-19 deaths, thus hampering a detailed analysis about the drivers of the second wave. However, already during the course of the first wave important factors had been identified internationally and also in Germany.

### SOCIAL DISPARITIES

There is a general consensus that infections and deaths have hit lower social groups the hardest, mainly due to higher mobility during the pandemic (Wachtler et al., 2020) and differences in the capacity for social distancing (Weill et al., 2020). Swedish registry data suggested that the most disadvantaged members of society suffered the most from SARS-CoV-2 infections and COVID-19 deaths (Drefahl et al., 2020).

One of the few individual-level studies for Germany based on health claims data, showed that the risk of contracting COVID-19 was highest in occupations where workers had frequent face-to-face contact with COVID-19 patients or potentially infected individuals during their occupational activities. However, increased risks were also observed in occupations with cramped workplaces, suboptimal hygienic condition, and among persons with temporary employment through agencies (Möhner and Wolik, 2020). A further study based on health claims data suggested strong social disparities in hospitalizations due to COVID-19 (Wahrendorf et al., 2021).

German ecological studies correlating regional characteristics with SARS-CoV-2 infections and COVID-19 deaths yielded contradictory results. Plümper & Neumayer (2020) concluded that only in the second phase of the pandemic and controlling for temporal dependence were predominantly poorer counties affected by COVID-19, whereas Wachtler et al. (2020) described a change in the social distribution of infections from regions with high to those with low social status already during the first wave. These studies used a small number of preselected indicators to characterize the regions. A different approach was taken by two studies (Doblhammer et al., 2020; Scarpone et al., 2020) that relied on machine learning algorithms to identify key regional features that predict SARS-CoV-2 infections and COVID-19 deaths without selecting specific indicators. They concluded that social status played an important role, in addition to features related to geographic location, hotspot events associated with the southern German carnival season, and the proportion of the population at risk in nursing homes. And they showed that the social gradient already evolved from a positive to a negative one during the first wave (Doblhammer et al., 2020).

### ETHNIC MINORITIES AND MIGRANTS

U.S. and U.K. studies were the first and most prominent to indicate that persons of black and Asian ethnicity were at increased risk for COVID-19 compared with whites (Sze et al., 2020), as were Native American and Latin communities (Tai et al., 2021). In Spain, migrants from sub-Saharan Africa, the Caribbean, and Latin America had an increased risk compared with Hispanics or migrants from Europe, North Africa, or Asia (Guijarro et al., 2021). Although minority groups are disproportionately affected by chronic disease, cardio-metabolic factors, and low 25(OH)-vitamin D levels, this cannot explain all of the disadvantage (Kirby, 2020). Poorer access to health care and disadvantaged living and working conditions appear to play an important role. With the exception of Doblhammer et al. (2020), who found that the proportion of foreigners and migrants was an important regional characteristic for COVID-19 in Germany during the first wave, we are not aware of any other studies for Germany. This is all the more remarkable as the studies above suggested that working environments, cramped living conditions, and language barriers are at odds with social distancing and should put this groups at higher risk.

### VALUES, NORMS AND COMPLIANCE

While social distancing is essential to contain the spread of COVID-19, not everyone is willing to comply with social distancing measures. From the U.S., there are reports based on debit card transaction data that Democrats are more likely to switch to remote spending after implementing government orders (Painter and Qiu, 2020). Also for the U.S., political conservatism inversely predicted compliance with behaviours aimed at preventing the spread of COVID-19 (Müller and Rau, 2021). For Germany, the “Institute for Democracy and Civil Society” found a correlation of COVID-19 illnesses and deaths with the proportion of voters for the party “Alternative for Germany (AfD)” in the last federal election (Medical Tribune, 2021). Individuals from this party sometimes take prominent roles in protests against Corona measures (Salheiser and Richter, 2020). However, regions with a high proportion of AfD voters border the neighbouring country of the Czech Republic, a hotspot of infection rates, so it is possible that infections have spilled over from there.

The aim of this study was to identify the key features explaining SARS-CoV-2 infections and COVID-19 deaths during the upswing of the second wave in Germany. We distinguished five two-week periods, beginning with low infection numbers from Oct. 1-15, 2020. We continued with the period Oct. 16-31, 2020, when the exponential increase in infections developed. The third period extends from Nov 1-15, 2020, and the fourth from Nov 16-30, 2020. In both periods exponential increase continued. The final period, from Dec 1-15, 2020, was characterized by the continued increase in infections, despite the onset of the “lockdown light”. The peak of infections was reached around Christmas time, which is outside our study period. We did not preselect regional characteristics but included a large number of maximally diverse indicators. Using machine learning and a framework for interpreting predictions, we identified the most important features and their association with regional SARS-CoV-2 infections and COVID-19 deaths.

## DATA AND METHODS

### DATA

We downloaded data (26 January 2021) from the Robert Koch-Institute (Robert Koch Institute and ESRI) which provides information on COVID-19 diagnoses and deaths by sex, age (age groups: 0-4, 5-14, 15-34, 35-59, 60-79, 80+), and county (NUTS3 region). Patients were not involved in this study.

Population size on county level was derived from the DESTATIS regional database at the end of the year 2019 (Statistical Offices of the Federation and the Länder, 2021). We calculated directly age-standardized incidence and death rates on the county level to control for differences in the age-distributions. We used the German age distribution from the year 2019 from the Regional database of the Statistical Offices of the Federation and the Länder (2021).

Macro variables characterized counties in the following domains (number of indicators in brackets)

1. SES-socioeconomic status (61),
2. Urbanity/density (22),
3. Health (20)
4. Care need (7),
5. Regional connectedness (15),
6. Norms and values (3),
7. Special geographic location (11),
8. Population composition in terms of foreigners/people with migration background (4),
9. Ageing and (age) structure of the population (12).

The data stem from the INKAR (Indikatoren und Karten zur Raumund Stadtentwicklung) database (2020) of the Federal Institute for Research on Building, Urban Affairs and Spatial Development (BBSR) (Federal Institute for Research on Building, Urban Affairs, and Spatial Development, 2020), latitude and longitude were defined in terms of the centres of the county capitals. The proportion of Catholics stems from the 2011 census, the PM10 emission data from the German Environment Agency Database (UBA), main diagnoses in hospitals by place of residence in 2017 from the Regional database of the Statistical Offices of the Federation and the Länder (2021), and the international COVID-19 incidence rates from the European Center for Disease Prevention and Control (2020). See Data Availability Statement below for access to the data and Table 1 in Suppl. Material for the list of independent variables and their descriptives. All 155indicators are numeric or dummies taking the values zero or one.

**Table 1:** Distribution of age-standardized COVID-19 incidence and death rates per 100,000 person-years by period (n=401 counties, IQR interquartile range)

| Period | Mean | SD | Min | 10% | 25% | 50% | 75% | 90% | Max | IQR |
| --- | --- | --- | --- | --- | --- | --- | --- | --- | --- | --- |
| Incidence rate per 100,000 population (age-standardized) |  |  |  |  |  |  |  |  |  |  |
| 01.-15.10. | 52.45 | 36.79 | 3.43 | 15.07 | 26.03 | 44.78 | 67.13 | 104.67 | 267.67 | 41.10 |
| 16.-31.10. | 196.86 | 100.08 | 21.63 | 82.17 | 124.02 | 180.95 | 250.00 | 331.38 | 613.03 | 125.98 |
| 01.-15.11. | 294.75 | 132.15 | 51.45 | 128.84 | 187.58 | 295.15 | 375.24 | 456.30 | 734.39 | 187.66 |
| 16.-30.11. | 299.44 | 144.25 | 20.30 | 135.46 | 203.50 | 286.18 | 371.91 | 481.42 | 968.59 | 168.41 |
| 01.-15.12. | 360.45 | 194.99 | 35.50 | 176.19 | 245.80 | 322.59 | 428.30 | 564.44 | 1239.70 | 182.49 |
| Death rate per 100,000 population (age-standardized) |  |  |  |  |  |  |  |  |  |  |
| 01.-15.10. | 0.50 | 0.89 | 0.00 | 0.00 | 0.00 | 0.00 | 0.74 | 1.55 | 4.68 | 0.74 |
| 16.-31.10. | 2.46 | 2.77 | 0.00 | 0.00 | 0.48 | 1.70 | 3.26 | 6.18 | 15.95 | 2.78 |
| 01.-15.11. | 4.85 | 4.09 | 0.00 | 0.81 | 1.91 | 3.78 | 6.79 | 10.34 | 29.78 | 4.88 |
| 16.-30.11. | 7.96 | 7.88 | 0.00 | 1.24 | 3.02 | 6.11 | 10.70 | 15.69 | 64.76 | 7.68 |
| 01.-15.12. | 11.66 | 9.76 | 0.00 | 2.67 | 5.24 | 8.94 | 15.34 | 24.98 | 68.45 | 10.10 |

### ANALYSIS STRATEGY

To explore the importance and the direction of the correlation of the regional correlates with age-standardized COVID-19 incidence and deaths we used the SHAP (SHapley Additive exPlanations) procedure (Lundberg and Lee, 2017). Our analysis strategy consisted of two steps: First, we trained gradient boosting models to predict for each period the age-standardized incidence and death rates with the 155 characteristics of the counties; these characteristics are termed features (Figure 1) and consist of all variables of the nine domains described above. The models also included the previous period’s age-standardized incidence rates. For counties bordering neighbouring countries, the previous period’s age-standardized incidence in the neighbouring country was also included. Gradient boosting models were trained using the CatBoostRegressor from the CatBoost algorithm (Prokhorenkova et al., 2018). As an alternative, we used the random forest regressor from the Scikit-learn module in Python (Pedregosa et al., 2011) with 5,000 trees. We kept all other hyperparameters at their default values. We calculated the R^2^ and root mean squared (RMSE) errors to evaluate how well the models fit the data. Second, we used Shapley values to characterize the 20 most prominent features in terms of negative/positive correlations with each of the two outcome variables. We displayed the Shapley values in summary plots in the Suppl. material. Each point on the summary plot is a Shapley value for a feature and a county. The position on the y-axis is determined by the feature and on the x-axis by the Shapley value. The colour represents the value of the feature from low to high. Overlapping points are jittered in y-axis direction, to get a sense of the distribution of the Shapley values per feature. The features are ordered according to their importance.

**Figure 1:**
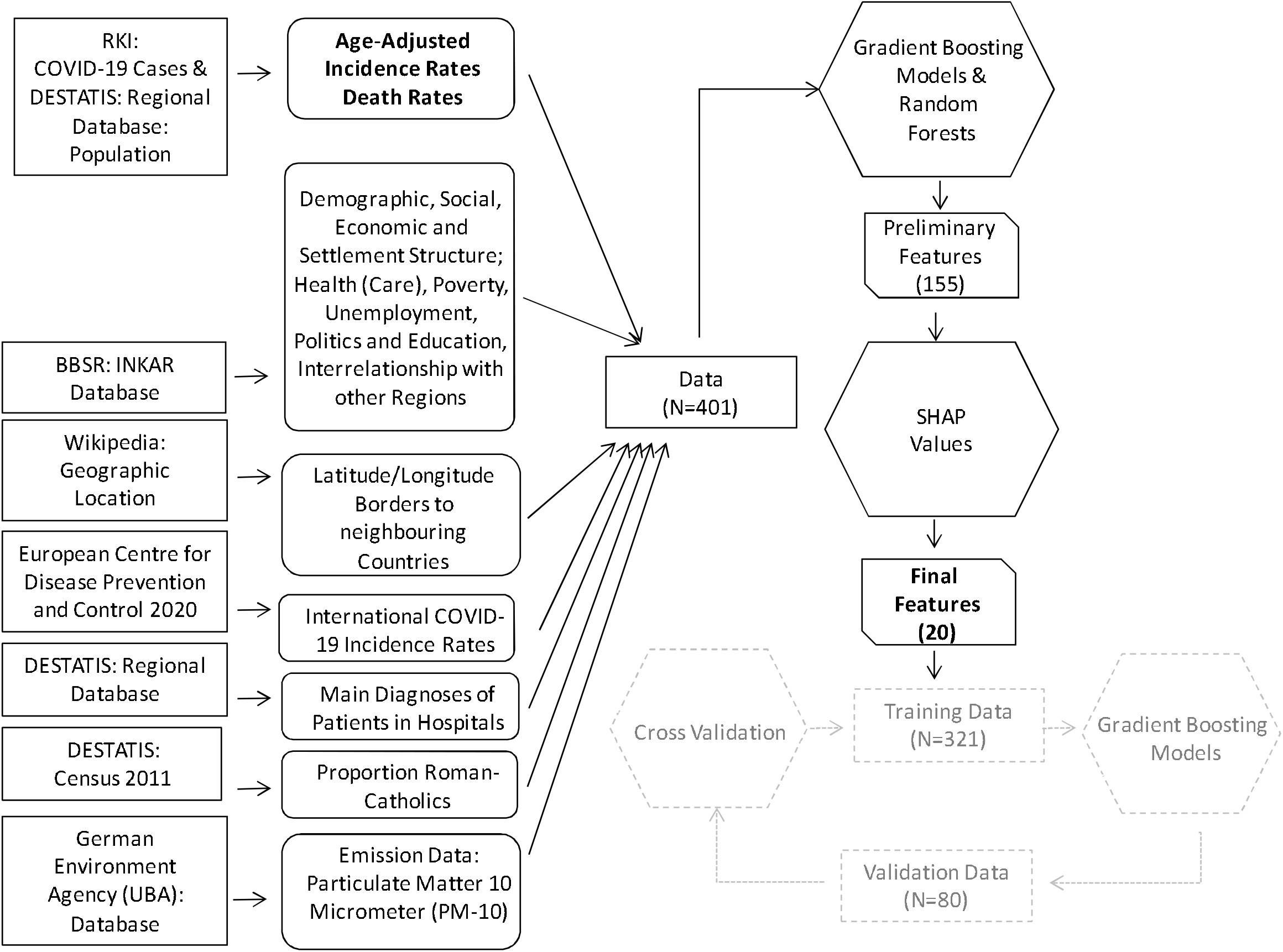
Analysis Strategy

We categorized the top 20 associations identified by the Shapley values into twelve categories depicting the correlation between the feature and the outcome: 1=positive SES gradient (SES high): higher incidence rates in high SES regions; 2=negative SES gradient (SES low): higher incidence in low SES regions; 3=urban/high density gradient (urban): higher incidence in urban/high density regions; 4=rural/low density gradient (rural): higher incidence in rural/low density regions; 5=care need gradient: higher incidence associated with high care need; 6= health gradient: higher incidence associated with poor health; 7= community’s connectedness low (connect low): higher incidence associated with low connectedness; 8=community’s connectedness high (connect high): higher incidence associated with high connectedness; 9=international migration high (migration high): higher incidence associated with higher proportion of foreigner/people with migration background; 10= geography; 11= values and norms; 12=age/aging structure of the region. We counted the number of features in each category and interpreted the evolution of their frequency over time, as well as the ranking of each feature.

As a sensitivity analysis, we repeated step one and fit a second model for each period using only the 20 most prominent features from the first model. We calculated R^2^ and RMSE to evaluate how much variance in the age-standardized incidence rates is covered by these 20 features.

To evaluate the out-of-sample model performance, we applied k-fold random subsampling (Berrar, 2019) using 20 folds. For each period we split the data at random to fit a model on a training set (80%) using the 20 most prominent features. This model was used to make predictions on a test set (20%) and to calculate the RMSE. Then a linear regression model was applied to explain the predictions by the actual response values from the test set. R^2^ from the linear regression model indicated how much variance from the actual response values can be explained by the predictions.

As an additional sensitivity analysis we identified all features with pairwise correlations smaller/larger than −0.8/+0.8 and excluded one of the features. All analyses were performed using Python 3.8.3.

## RESULTS

### AGE STANDARDIZED COVID-19 INCIDENCE AND DEATH RATES IN THE FIVE PERIODS

Age-standardized incidence rates (Table 1) quadrupled from the first half of October (October 1-15: 52.45 per 100,000) to the second half (196.86), reaching 300 cases in the second half of November and 360 in the first half of December.

Age-standardized death rates (Table 1) were still low in early October (October 1-15: 0.50 new cases per 100,000), then doubled almost every two weeks from mid-October onward (October 16-31: 2.46) and reached a maximum in December (December 1-15: 11.66).

These growth curves showed distinct geographic patterns that changed over time, as shown in Suppl. Material Figures 1-5. Both incidence and mortality moved from west to east, starting in the high-incidence regions of North Rhine-Westphalia, Baden-Württemberg, and Bavaria and moving to Saxony, Thuringia, and western Bavaria.

### MODEL FITTING AND DIAGNOSTICS

Boosting models generally outperformed random forests in terms of accuracy therefore we have only shown the results of the boosting models (the results of the random forests are available on request). A sensitivity analysis using only the 20 most important features to fit the boosting models resulted in almost unchanged R^2^ values but increased RMSE values (Supplementary Material Tables 2). This indicates that the boosting algorithm produces well-fitted models even when only a subset of the most prominent features was used. Out-of-sample performance increased across periods, with the performance of the models for death rates always well below that for incidence rates (incidence rate: R^2^=0.4911 in the first period to 0.7428 in the last period; death rate: R^2^=0.1722 in the first period to R^2^=0.4213 in the last period). The descriptive statistics of the outcome variables are shown in Table 1 and the twenty most important features are shown, among others, in Table 1 in the Suppl. Material.

**Table 2:** Number of features in the top 10/20 features indicating a positive correlation with high COVID-19 incidence and death rates

| Correlation with high incidence | 1-15 Oct |  |  |  | 16-31 Oct |  |  |  | 1-15 Nov |  |  |  | 16-30 Nov |  |  |  | 1-15 Dec |  |  |  |
| --- | --- | --- | --- | --- | --- | --- | --- | --- | --- | --- | --- | --- | --- | --- | --- | --- | --- | --- | --- | --- |
|  | IR 10 | IR 20 | DR 10 | DR 20 | IR 10 | IR 20 | DR 10 | DR 20 | IR 10 | IR 20 | DR 10 | DR 20 | IR 10 | IR 20 | DR 10 | DR 20 | IR 10 | IR 20 | DR 10 | DR 20 |
| SES high (1)* | 2 | 3 | 2 | 3 | 0 | 0 | 1 | 1 | 0 | 0 | 2 | 4 | 0 | 1 | 0 | 3 | 2 | 3 | 3 | 3 |
| SES low (2) | 0 | 3 | 1 | 4 | 4 | 8 | 3 | 4 | 2 | 5 | 1 | 5 | 2 | 5 | 4 | 7 | 3 | 4 | 1 | 1 |
| Urban/high density (3) | 1 | 3 | 2 | 3 | 0 | 1 | 1 | 2 | 0 | 2 | 0 | 1 | 0 | 0 | 1 | 2 | 0 | 1 | 1 | 4 |
| Rural/low density (4) | 1 | 1 | 1 | 2 | 2 | 2 | 1 | 2 | 1 | 2 | 0 | 0 | 2 | 2 | 0 | 1 | 1 | 1 | 0 | 1 |
| Care need (5) | 0 | 1 | 0 | 2 | 0 | 0 | 0 | 1 | 0 | 0 | 0 | 0 | 1 | 2 | 1 | 2 | 1 | 2 | 0 | 0 |
| Health (6) | 1 | 2 | 1 | 2 | 1 | 1 | 1 | 2 | 1 | 1 | 2 | 2 | 2 | 2 | 1 | 2 | 1 | 1 | 2 | 2 |
| Connect low (7) | 0 | 0 | 0 | 1 | 0 | 1 | 1 | 1 | 1 | 2 | 1 | 2 | 0 | 1 | 0 | 0 | 0 | 0 | 0 | 0 |
| Connect high (8) | 1 | 1 | 0 | 0 | 0 | 3 | 0 | 0 | 0 | 0 | 0 | 0 | 0 | 0 | 0 | 0 | 0 | 3 | 0 | 2 |
| Migration high (9) | 2 | 2 | 1 | 1 | 1 | 1 | 0 | 1 | 0 | 0 | 1 | 1 | 0 | 1 | 0 | 0 | 0 | 0 | 0 | 0 |
| Geography (10) | 1 | 2 | 1 | 1 | 1 | 2 | 1 | 2 | 2 | 2 | 1 | 1 | 2 | 2 | 2 | 2 | 1 | 2 | 1 | 2 |
| Pop. Char.: Values, norms (11) | 1 | 1 | 0 | 0 | 1 | 1 | 0 | 1 | 1 | 2 | 1 | 1 | 1 | 1 | 1 | 1 | 1 | 2 | 1 | 1 |
| Pop. Char.: Age, aging (12) | 0 | 1 | 1 | 1 | 0 | 0 | 1 | 3 | 2 | 4 | 1 | 3 | 0 | 3 | 0 | 0 | 0 | 1 | 1 | 4 |
| Total | 10 | 20 | 10 | 20 | 10 | 20 | 10 | 20 | 10 | 20 | 10 | 20 | 10 | 20 | 10 | 20 | 10 | 20 | 10 | 20 |

IR 10: Incidence rate first 10 features; IR 20: Incidence rate first 20 features
DR 10: Death rate first 10 features; DR 20: Death rate first 20 features
\*: 1=positive SES gradient (SES high); 2=negative SES gradient (SES low); 3=urban/high density gradient (urban); 4=rural/low density gradient (rural); 5=care need stationary/ambulant; 6=health; 7= positive gradient with community's connectedness low (connect low); 8=positive gradient with community's connectedness high (connect high); 9= positive gradient with international migration high (migration high); 10= Geography; 11= population characteristics: Values & norms; 12=population characteristics: Age, aging.

### MODEL RESULTS

The age-standardized incidence and death rates changed over time, as did the key features. Taking the first twenty features according to their Shapley values and grouping them into the categories outlined above, we find that features related to SES, urbanity/density, and health were present in all time periods; those representing the connectedness of a region were important in the period from mid-October to mid-November and again in December. Features related to need for care gained importance in the second half of November and in December, while those related to migration were important in October and the first half of November. Features reflecting values and norms gained importance over time, as did those characterizing the (age) structure and aging process in a region (Table 2).

#### Socioeconomic Status

At the beginning of the second wave (Oct-1-15), the overall low incidence was comparatively elevated in both high and low SES regions (Figure 2a: of the first 20 features, three were associated with low SES and three with high SES); with the exponential increase in infections, low SES regions became the driving forces; no single feature was associated with high SES (Figure 2a: 6. Oct 16-31 (8 of 20 features related to low SES); Nov-1-15 (5 of 20 features related to low SES). During the peak period (Nov-16-30; Dec-1-15), infections again spilled over from low to high SES regions (Figure 2a).

**Figure 2:**
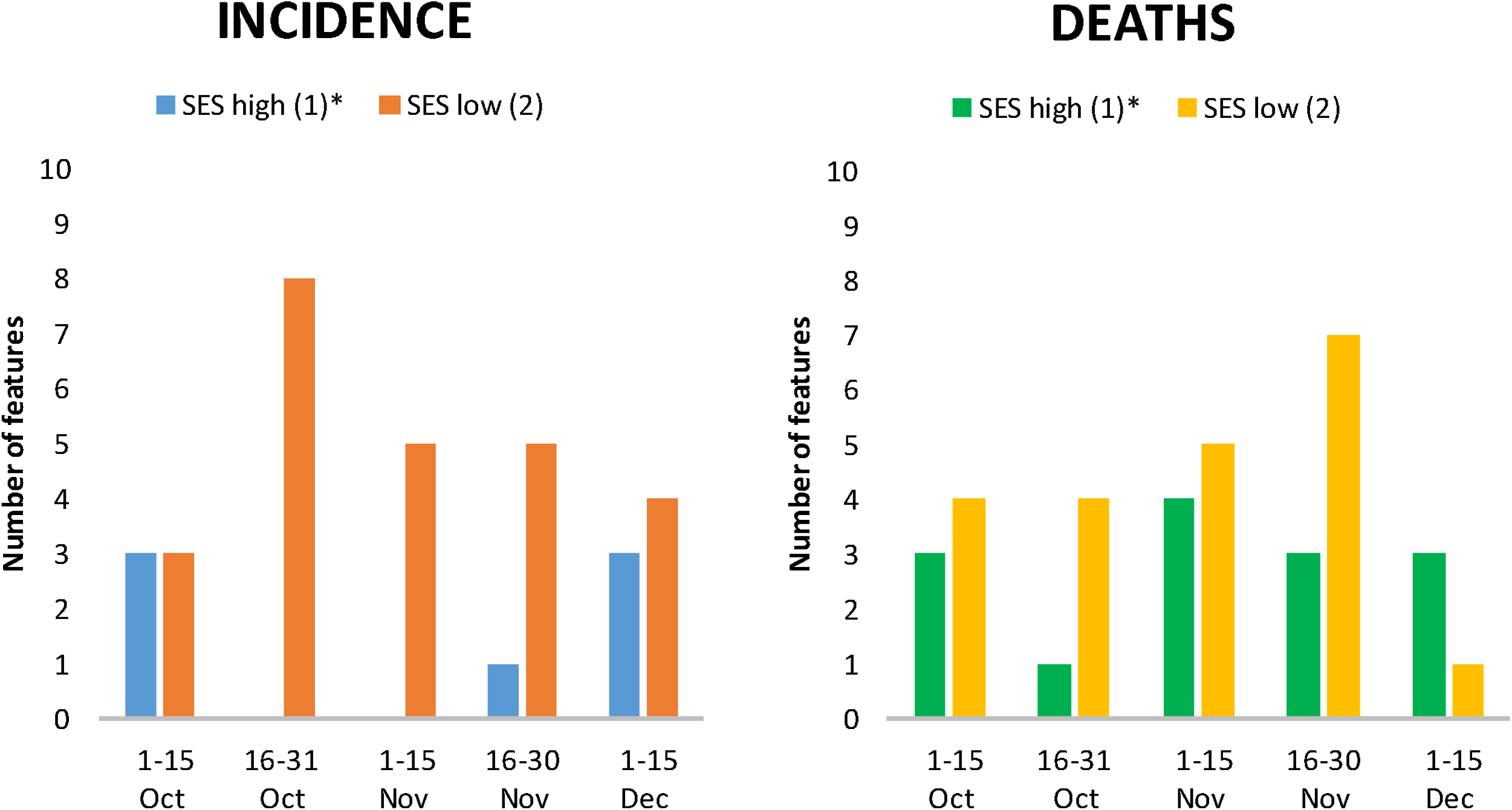
Number of features in the top 20 showing the relationship between low and high SES, and incidence (a) and death rates (b), by time period.

COVID-19 deaths were correlated with both low and high SES regions in all periods, with generally higher correlation with low SES regions (Figure 2b).

Of note, at the beginning of the second wave, features that indicated a positive correlation of infections and deaths with high SES regions ranked ahead of those that correlated with low SES regions (Table 2). This changed in the second period. For example, in the second period, the top ranking SES features for both infections and deaths indicated a negative gradient (infections rank 6: “%people without a vocational qualification in all employed persons in 2017”; mortality rank 2: “%change in long-term unemployment rate in 2012-2017”) (Suppl. Material Figures 6-10).

#### Migration

Features related to international migration background were among the top characteristics until mid-to-late November and correlated positively with incidence (Figure 3a) and death rates (Figure 3b). In the first period, the regional “%proportion of foreigners” ranked third in incidence and ninth in mortality. In the second period, it still ranked fourth in incidence and eleventh in mortality, and in the third period, it ranked sixth in mortality (Suppl. Material Figures 6,7,8).

**Figure 3:**
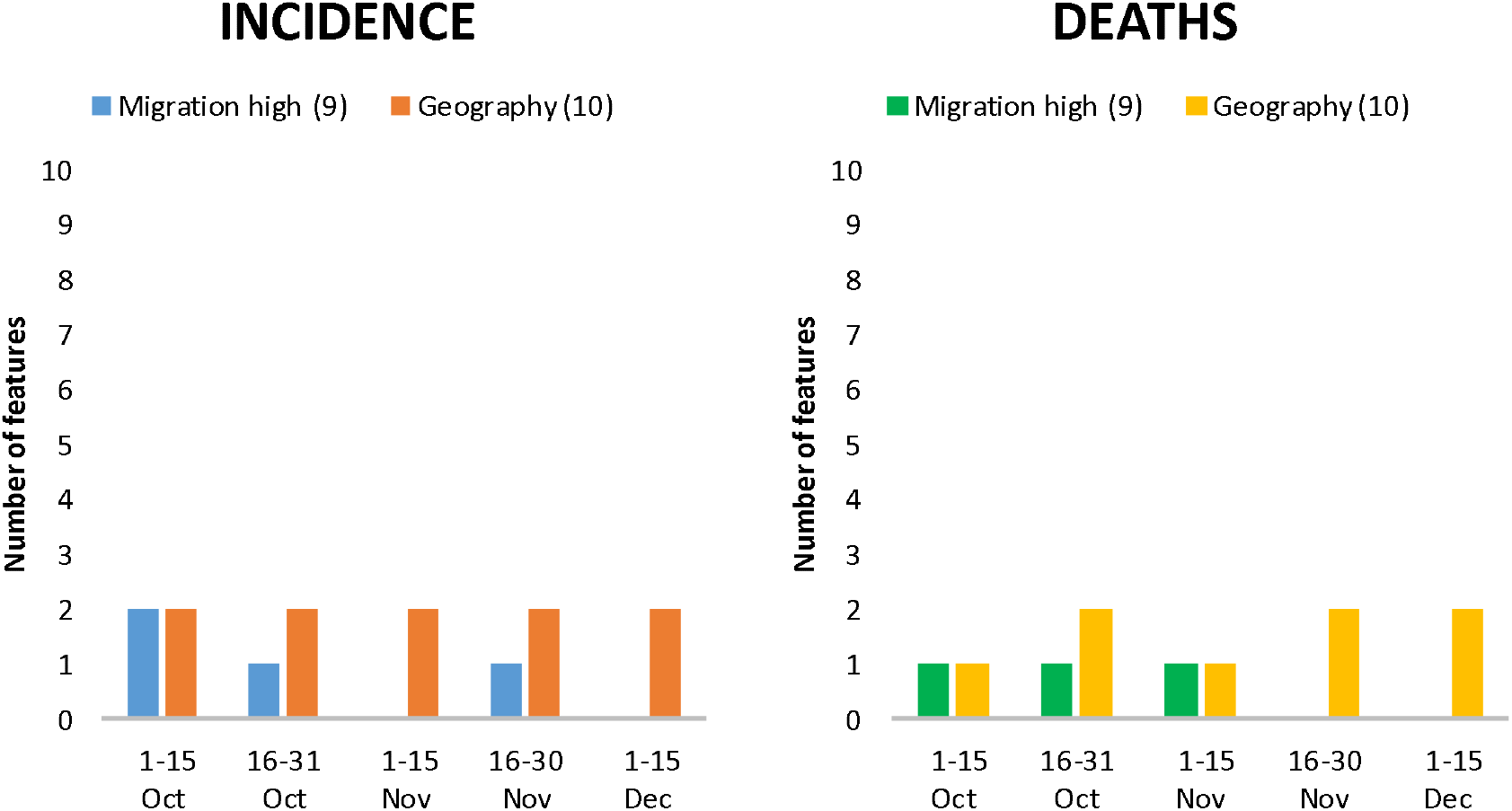
Number of features in the top 20 showing the relationship between migration and geographic dispersion, and incidence (a) and death rates (b), by time period.

#### Geography, urbanity/rurality, connectedness

Geography in the form of latitude and longitude, which can be considered a residual category in characterizing the disease spread, was among the top features in all periods (Figures 3a and 3b). In the second period, the characteristics “Cum. Incidence per 100,000 for 14 days in neighbouring country on day 289” and “Federal border with Austria” and their positive correlation with incidence (Suppl. Material Figure 7) indicated a spill-over effect of the high incidence in Austria to Germany. Geographical spread was independent of urbanity/rurality (Figure 4a and 4b) and connectivity of a region (Figure 5a and 5b). During the wave, infections and deaths occurred in both urban and rural regions, and in better or less connected regions. It is interesting to note the positive correlation of “Nitrogen surplus per agricultural area in kg/ha in 2016” (rank 2) and “Average PM10” with deaths (rank 4) in the first period (Suppl. Material Figure 6).

**Figure 4:**
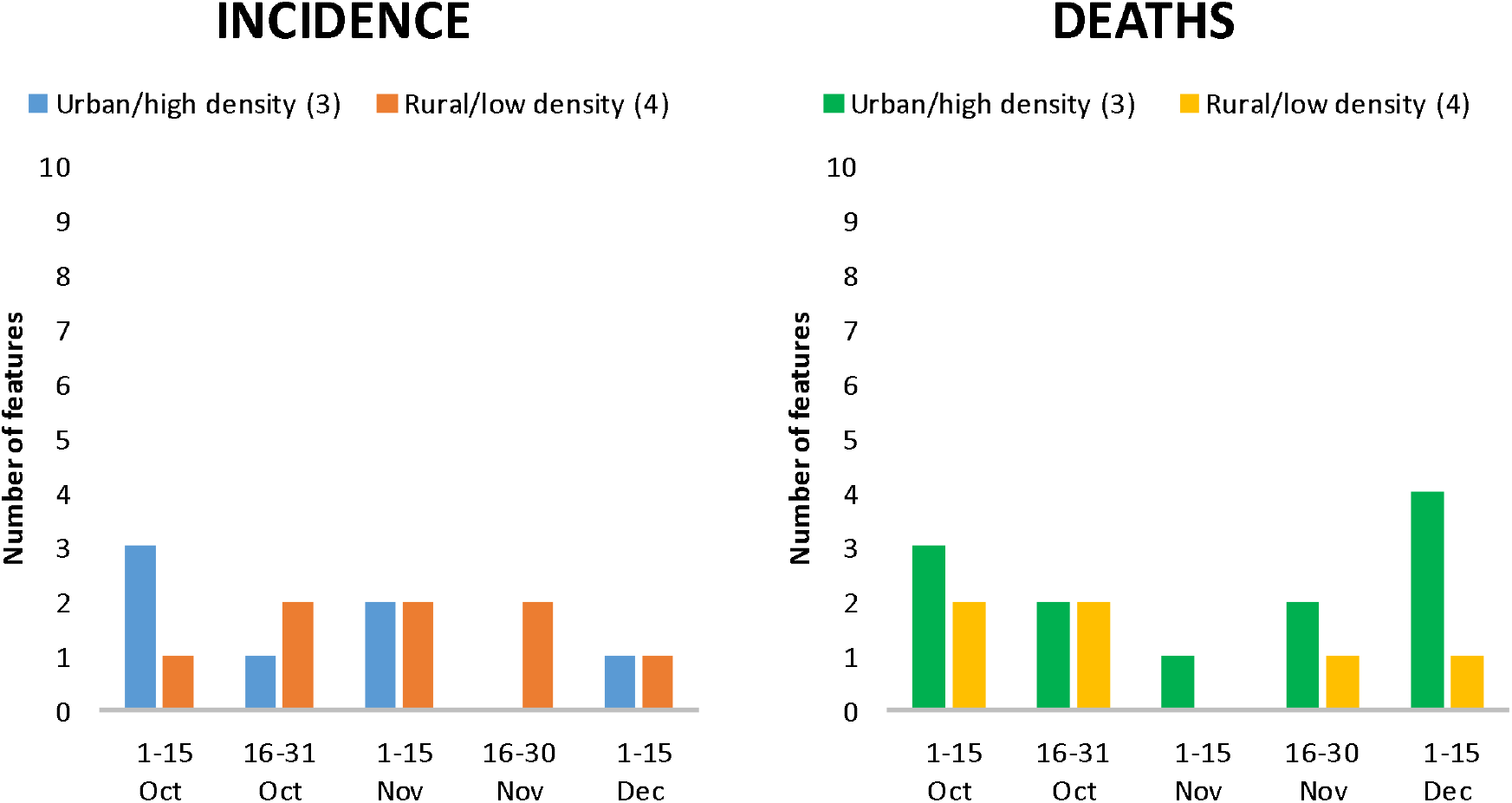
Number of features in the top 20 showing the relationship between urbanity/rurality and density, and incidence (a) and death rates (b), by time period.

**Figure 5:**
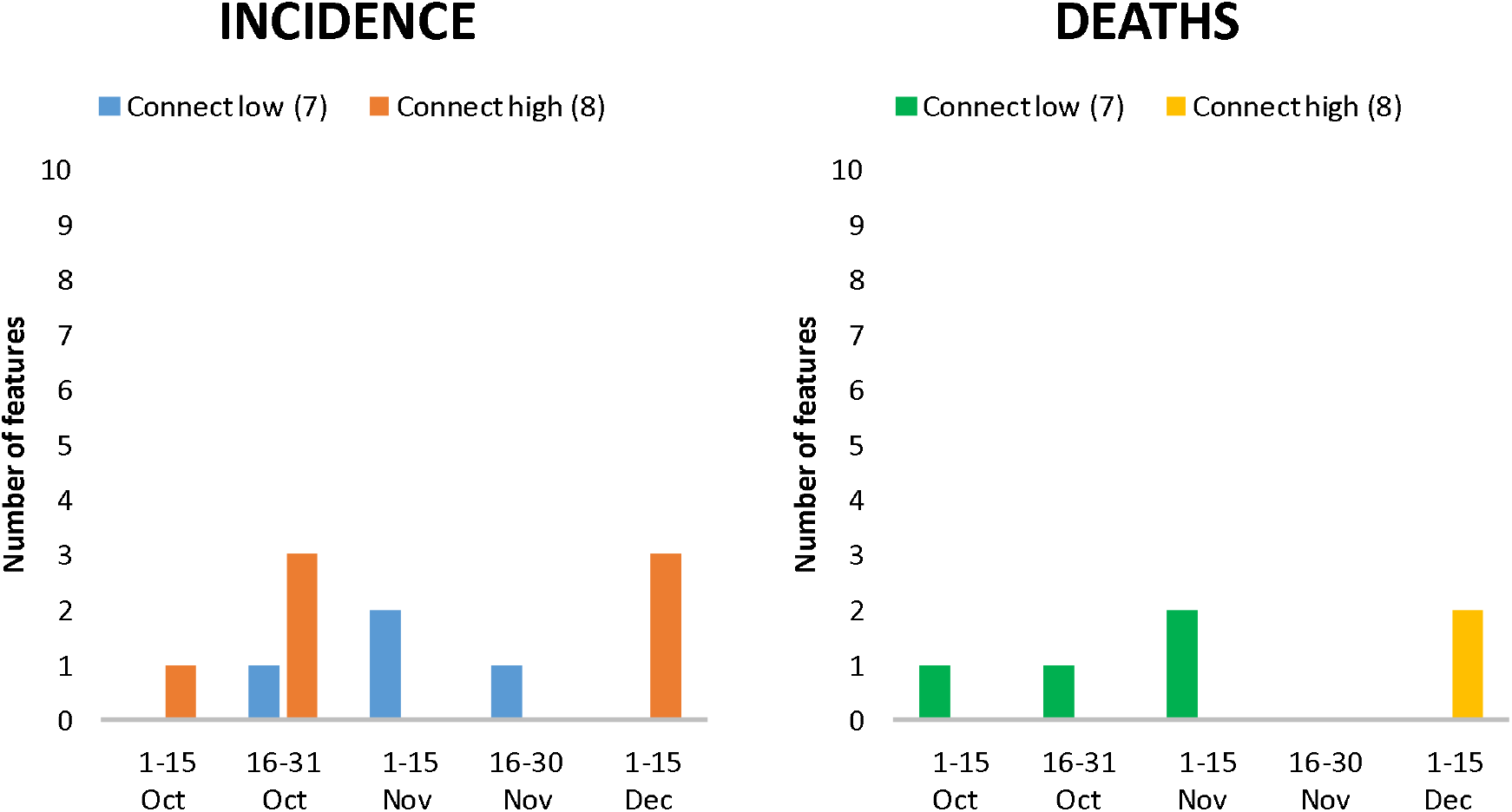
Number of features in the top 20 showing the relationship between connectedness, and incidence (a) and death rates (b), by time period.

**Figure 6:**
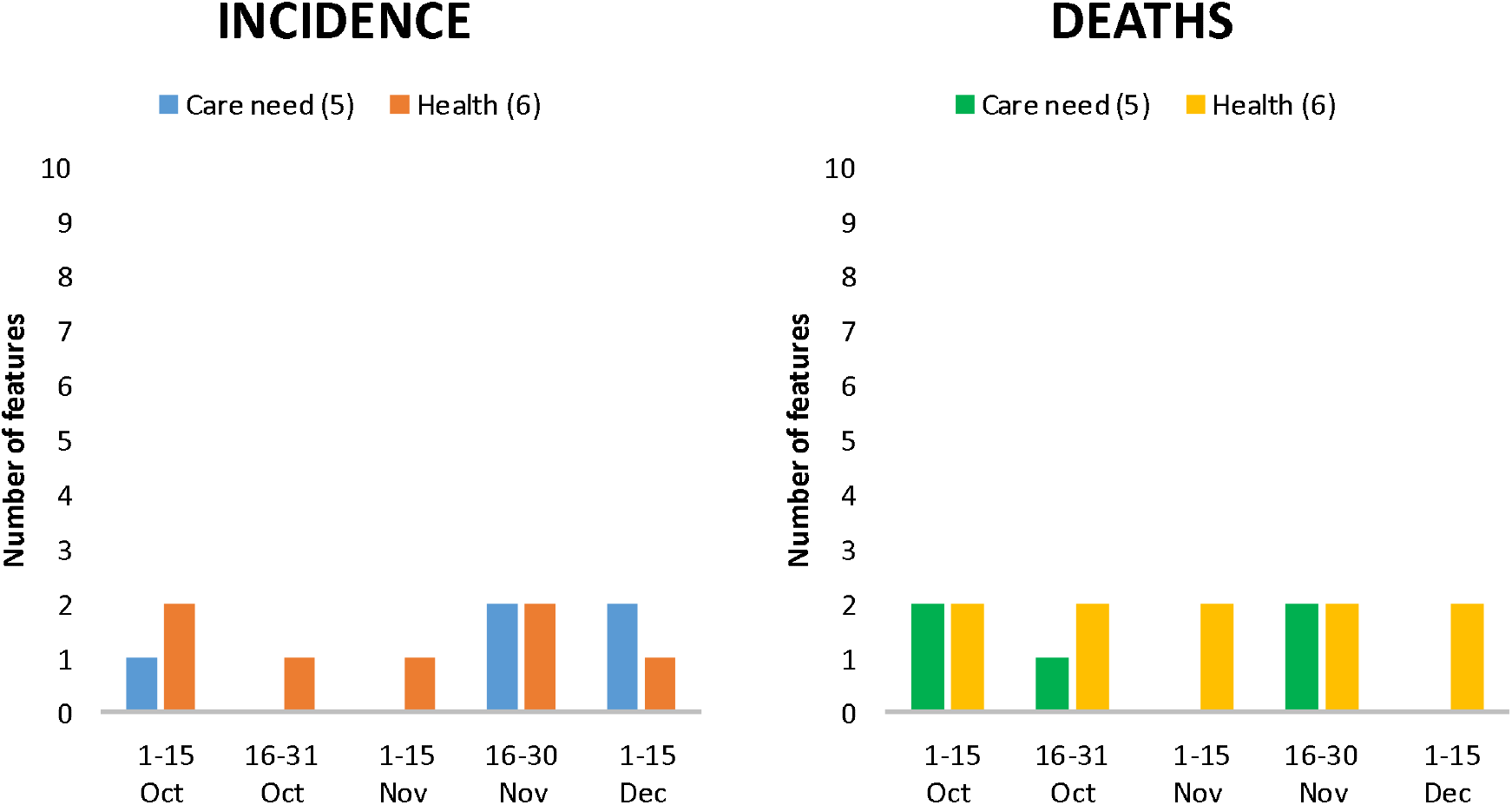
Number of features in the top 20 showing the relationship between care need and health, and incidence (a) and death rates (b), by time period.

**Figure 7:**
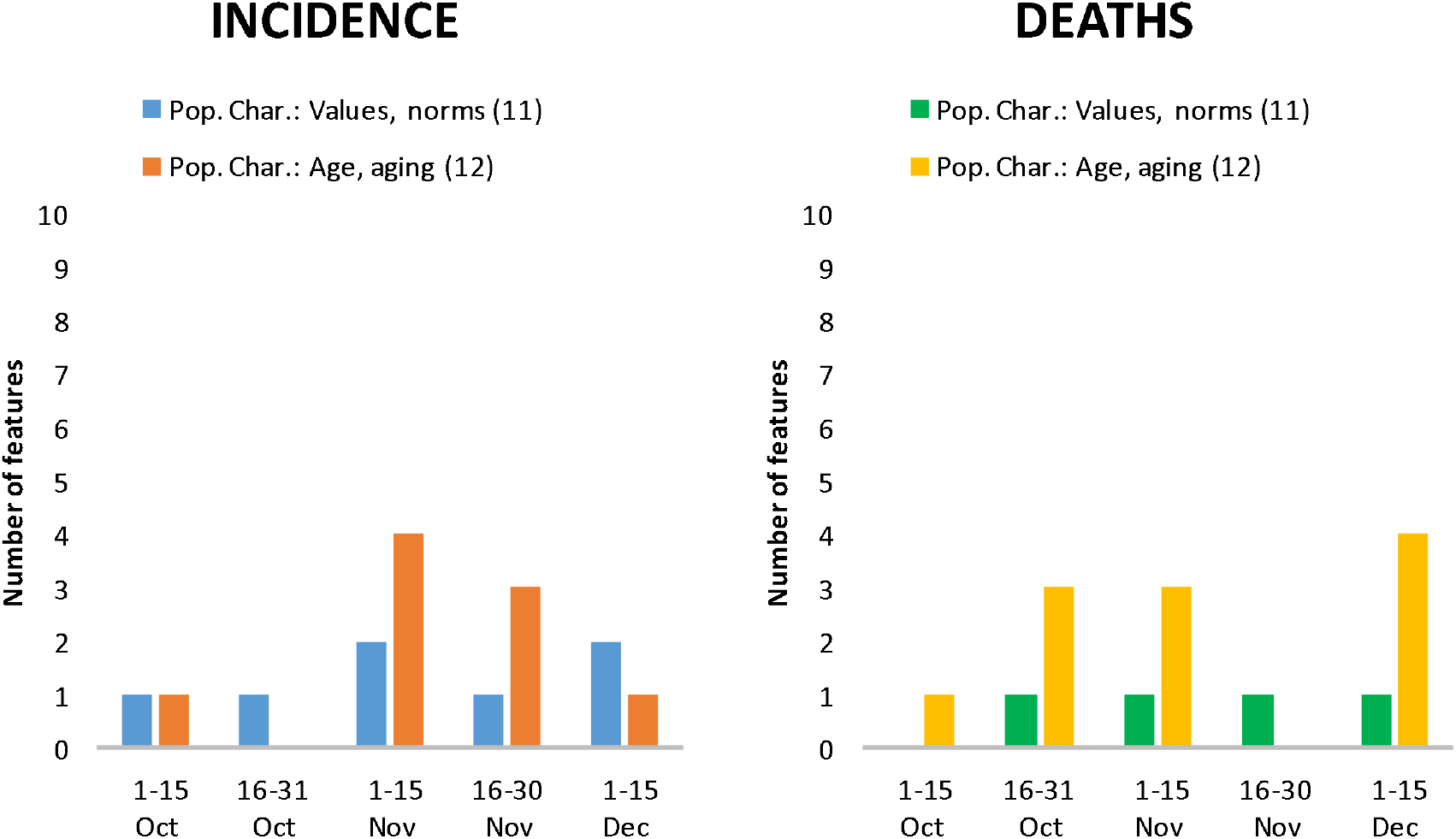
Number of features in the top 20 showing the relationship between values/norms and age/ageing, and incidence (a) and death rates (b), by time period.

#### Care need and health

The proportion of the population in need of long-term care is another important characteristic, as suggested by its presence among the top 20 in all periods (Figures 6a and 6b). In the first period, the feature “%persons in long-term inpatient care out of all persons in need of care” was among the top 20 but negatively correlated with incidence and death rates, suggesting fewer infections and lower mortality for regions with a high proportion of the population in need of care. This changed in the second period, when “number of beds in nursing homes” was positively correlated with death rates. In the fourth period, the feature “%persons in outpatient care among all persons in need of long-term care” and “use of long-term care services per 10,000 persons in 2017”, which refer to persons not living in nursing homes, were negatively correlated with incidence and death rates, again indicating a higher risk of regions with a high proportion of the population living in nursing homes (Suppl. Material Figures 6,7,9).

Characteristics representing population health (Figures 6a and 6b) were generally negatively correlated with incidence and death rates. The highest-ranking feature in all periods was the incidence of SARS-CoV-2 infections in the previous period (Suppl. Material Figures 6-10). However, there was also a positive correlation between prior-period infections and deaths and the proportion of persons with a diagnosis of endocrine, nutritional, and metabolic diseases, which includes diabetes (periods 3 and 4, Suppl. Material Figures 8 and 9).

#### Values and Norms, and Age/Ageing

Of the features that depict values and norms (Figures 7a and 7b), one consistently appeared in the top 20, was positively correlated with incidence and death rates, and increased in importance over the course of the wave. In the second period, the feature “%Valid votes for AfD out of all valid votes in 2017” ranked 13th (deaths only), in the third period it ranked 8th (incidence) and 7th (deaths), in the fourth period it ranked 3rd (incidence) and 2nd (deaths), and in the fifth period it ranked 3rd (incidence) and 5th (deaths) (Suppl. Material, Figures 6-10). Finally, population characteristics related to age and (aging) structure (Figures 7a and 7b) played an important role with decreasing importance for incidence but increasing one for deaths over the period.

## DISCUSSION

Combining an ecological study design with machine learning techniques using 155 county-level regional variables, we examined potential associations between regional characteristics and SARS-CoV-2 infections and COVID-19 deaths. This study design helped us to avoid imposing our expectations on the pre-selection of possible regional characteristics. We restricted out analysis to the period between beginning of October and mid-December 2020, defined as the second-wave upswing (Schilling et al., 2021), and differentiated five two-week periods. These periods reflect exponentially increasing infections which peaked in mid-December, followed by increases in deaths.

Restricting our analysis to those first twenty risk factors identified by Shapley values, we concluded that, similar to the first wave (Doblhammer et al., 2020), SES was an important driver in the second wave. While both social gradients, positive and negative, were present in SARS-CoV-2 infections in October, the negative SES gradient began to dominate over time and was always the dominant one in mortality. Higher mobility of high SES groups in periods of low infection and a greater decrease in mobility in periods of high infection may explain this trend. US studies showed that the poorest areas moved from lowest to highest mobility and thus had fewer opportunities for social distancing (Weill et al., 2020).

International migration and a high proportion of foreigners living in a county were important regional characteristics during those periods when the exponential increase in incidence intensified. On the one hand, in these regions, international contacts may have been higher and more infections may have been imported from abroad. For the beginning of the exponential increase (second half of October), we can show spill-over effects of infections from the neighbouring country Austria and, in general, from neighbouring countries with high infection rates. On the other hand, a high proportion of foreigners may be indicative of a negative social gradient, as they often work in occupations such as cleaners, food production workers, or elderly care (Federal Statistical Office, 2020), and are therefore more exposed to the virus. A high proportion of foreigners may also be related to the presence of different cultures and norms, a lack of access to information about the pandemic, and differences in compliance with protective measures such as wearing masks and maintaining social distance. On the one hand, results from the UCL COVID-19 Social Study suggested that working outside the home was associated with lower compliance (Wright et al., 2021). On the other hand, in Switzerland non-compliance with COVID-19-related public health measures was higher among young adults without migration background (Nivette et al., 2021).

In our study, characteristics of a region related to the nursing home population and care provided by ambulatory care services remained among the top features explaining high versus low rates of infection and death. This was despite the fact that, compared with the first wave, the proportion of COVID-19 deaths in nursing homes decreased as a proportion of all deaths (Ioannidis, Axfors, Contopoulos-Ioannidis 2021). The high importance of these features reflects the sad fact that this particularly vulnerable group is difficult to protect (Rothgang et al., 2020), despite commonly applied but also highly criticized isolation measures (Velayudhan et al., 2020; Morina et al., 2021). In addition, there was evidence that COVID-19 outbreaks in nursing homes were associated with spatial deprivation and that the latter was a major risk factor for COVID-19 deaths in nursing home residents (Bach-Mortensen and Degli Esposti, 2021).

Population health appeared to be another important characteristic which was associated with a negative SES gradient and influenced SARS-CoV-2 infections and COVID-19 mortality. We included information on regional health profiles reflecting known comorbidities of severe COVID-19 cases (Karagiannidis et al., 2020). Incidence and death rates were found to be higher in regions with a high proportion of diagnosed endocrine, nutritional, and metabolic diseases. This includes diabetes mellitus which has been repeatedly identified as a major risk factor for severe COVID-19 outcomes and mortality (Crankson et al., 2021).

One feature that became more important during the second wave was the proportion of the population that voted for the party “Alternative for Germany (AfD)” in the last federal election. In the exponential growth phase, it was the third most important characteristic of a region in terms of infections and the second most important in terms of deaths. Although this correlation has been noted before, it has also been highly controversial (Medical Tribune, 2021). We consider this characteristic as a possible indicator of compliance. Numerous surveys suggested that COVID-19 is a deeply partisan issue in the US and that partisanship was more strongly associated with physical disengagement than numerous other factors, including county SARS-CoV-2 infections, population density, median income, and racial and age demographics (Gollwitzer et al., 2020). Studies in Germany showed that respondents from Eastern Germany and those with little trust in public institutions were particularly critical of containment measures (Diehl and Wolter, 2020).

The “Deutschlandtrend” survey of the public broadcaster “ARD” found clear preferences among voters of the parties represented in the Bundestag. In mid-May 2020, 61 percent of AfD supporters were in favour of lifting containment measures, compared with 25 to 34 percent among supporters of the ruling coalition parties (infratest dimap, 2020). Note that the feature “%of Roman-Catholics “in a county, which was prominent in the first wave (Doblhammer et al., 2020), lost importance in the second wave. In the first wave, it reflected large gatherings during the carnival season in southern Germany that led to hotspots of infection (Streeck et al., 2020). During the second wave, large gatherings occurred mainly during demonstrations against Corona measures, and SARS-CoV-2 infection rates were particularly high in protesters’ origin regions after these demonstrations (Lange and Monscheuer, 2021).

Similar to the first wave, there was no pronounced urban/high density gradient or gradient associated with the connectedness of a region (Doblhammer et al., 2020). Population density per se did not appear to be a risk factor, which is supported by a regional analysis of COVID-19 prevalence in the United States (Paul et al., 2020) and Germany (Scarpone et al. 2020). Cities have both the healthiest and unhealthiest populations. The former benefit from better infrastructure and access to health care, while the latter have a higher burden of disease and lower life expectancy due to behavioural and environmental risk factors (Rydin et al., 2012).

### STUDY LIMITATIONS

Our study is hampered by a series of limitations. Reliance on the county level introduces the problem of the modifiable areal unit (Kirby et al., 2017). County-level data might be too course, but also too finely graded, to detect important features driving the pandemic. To overcome the limitation that the macro variables are restricted to Germany, we included the age standardized incidence in neighbouring countries for counties with international borders.

True infection rates are not known for SARS-CoV-19 due to asymptomatic individuals, regional approval criteria for testing that resulted in different testing rates and differences in reporting by local health departments to the RKI. In addition, these data report the time of diagnosis rather than the time of infection. There was also a strong weekday effect with lower reporting rates on weekends. Our 14-day period averages over these different lags to give an average picture of infections during this period. In addition, our models included information about infections in the previous period.

Different machine learning algorithms identify different features and their importance. We obtained similar results regardless of the machine learning algorithm used (Random Forests (results available upon request) versus Cat Boosting algorithms, with the latter better reflecting the data. Nevertheless, it is important to keep in mind that the interpreted Shapley values explain the model and not the data.

## CONCLUSION

Our study showed that an ecological study approach using explainable machine learning methods can help to shed more light on the infection pathways of COVID-19 in Germany. Ecological analyses have their place in stimulating innovation in a rapidly evolving field of research (Wu et al., 2020) where individual data are not yet available. Although ecological analysis cannot provide insight into mechanisms, it can highlight potential drivers. Our study showed that a bundle of factors was crucial for the increase in infections in the second wave. As in the first wave, there was spill-over of infection from high to low SES groups. Risky working conditions with reduced opportunities for social distancing, a high chronic disease burden, and residence in nursing homes were associated with the SES effect. In addition, noncompliance with Corona measures and spill-over effects from neighbouring countries were also found. To further support this finding, we urgently need more individual-level data (Khalatbari-Soltani et al., 2020).

## Data Availability

The following datasets were derived from sources in the public domain:
Robert Koch Institute, ESRI. RKI COVID19. dl-de/by-2-0. https://npgeo-corona-npgeo-de.hub.arcgis.com/datasets.
Statistical Offices of the Federation and the Laender. Regional database. https://www.regionalstatistik.de/genesis
DESTATIS Census 2011: Census database. https://ergebnisse.zensus2011.de
INKAR Database: Federal Institute for Research on Building, Urban Affairs, and Spatial Devel-opment. INKAR - Indikatoren und Karten zur Raum- und Stadtentwicklung 2020. https://www.inkar.de/
European Center for Disease Prevention and Control: Download historical data (to 14 De-cember 2020) on the daily number of new reported COVID-19 cases and deaths world-wide (https://www.ecdc.europa.eu/en/publications-data/download-todays-data-geographic-distribution-covid-19-cases-worldwide)
The following data are available on request from the data holder:
Emission data: German Environment Agency Database (UAB): https://www.umweltbundesamt.de/en

https://www.regionalstatistik.de/genesis

https://ergebnisse.zensus2011.de

https://www.inkar.de/

https://www.ecdc.europa.eu/en/publications-data/download-todays-data-geographic-distribution-covid-19-cases-worldwide

https://www.umweltbundesamt.de/en

