## Supplementary material for "The second wave of SARS-CoV-2 infections and COVID-19 deaths in Germany – driven by values, social status and migration background? A county-scale explainable machine learning approach"

Supplementary Table 1: List of independent variables and descriptives

| Variable name | Label | Category | Mean (sd) |
| --- | --- | --- | --- |
| D_Abhaen | Old-age (65+) dependency ratio in 2017 | Age/Aging | 34.35 (5.47) |
| D_Auslae | %Foreigners in total population in 2017 | Foreigners/Migration | 10.03 (5.15) |
| D_Aussen | International net migration per 1,000 persons in 2017 | Foreigners/Migration | 4.81 (6.63) |
| D_fertil | Total fertility rate in 2017 | Age/Aging | 1.63 (0.13) |
| D_Gesamt | Total net migration per 1,000 persons in 2017 | Foreigners/Migration | 5.1 (4.28) |
| D_ldergebur~201 | %Change of number of births in 1995-2017 | Age/Aging | 16.51 (8.78) |
| D_Lebens | Life expectancy at birth in 2017 | Health | 80.66 (1.01) |
| D_nerunter6ja~n | %Change of persons aged 5 and younger in all persons in 2012-2017 | Age/Aging | 11.58 (4.92) |
| D_nervon6bisu~r | %Change of persons aged 6 to <18 in all persons in 2012-2017 | Age/Aging | -1.1 (7.79) |
| D_nter18jah~201 | %Persons aged 6 to <18 in all persons in 2017 | Age/Aging | 10.8 (0.93) |
| D_unter6ja~2017 | %Persons aged 5 and younger in all persons in 2017 | Age/Aging | 5.37 (0.46) |
| D_v405 | %Change of number of persons at age 50-65 in 2012-2017 | Age/Aging | 9.54 (4.67) |
| D_v406 | %Change of number of persons at age 65-75 in 2012-2017 | Age/Aging | -3.95 (4.8) |
| D_v423 | Sex ratio (females to males) at age 20-40 in 2017 | Age/Aging | 0.92 (0.05) |
| D_v424 | %Change of number of persons in 2012-2017 | Age/Aging | 2.21 (2.78) |
| D_v425 | %Change of number of marriages in 2012-2017 | Age/Aging | 5.23 (9.95) |
| D_v483 | %Change of proportion of foreigners in 2012-2017 | Foreigners/Migration | 3.33 (1.11) |
| D_v599 | Remaining life expectancy at age 60 in 2017 | Health | 23.7 (.66) |
| D_Veraen | %Change of life expectancy at birth in 1993/94/95 and 2015/2016/2017 | Health | 7.49 (1.18) |
| flag_belgien | Border with Belgium | Geography | 0.01 (0.11) |
| flag_daenemark | Border with Denmark | Geography | 0.01 (0.09) |
| flag_frankreich | Border with France | Geography | 0.05 (0.21) |
| flag_luxemburg | Border with Luxemburg | Geography | 0.01 (0.1) |
| flag_niederla~e | Border with The Netherlands | Geography | 0.03 (0.18) |
| flag_oesterre~h | Border with Austria | Geography | 0.04 (0.2) |
| flag_polen | Border to Poland | Geography | 0.02 (0.15) |
| flag_schweiz | Border with Switzerland | Geography | 0.01 (0.11) |
| flag_tschechien | Border with Czech Republic | Geography | 0.04 (0.21) |
| G_latitude | Latitude | Geography | 50.62 (1.74) |
| G_longitude | Longitude | Geography | 9.87 (2.03) |
| HC_aerztever | General practitioner per 100,000 persons in 2017 | Health | 61.36 (26.11) |
| HC_Ambulante | %Persons in outpatient long-term care in all persons in long-term care in 2017 | Care need | 23.82 (5.23) |
| HC_Apotheken | Pharmacies per 100,000 persons in 2017 | Urbanity/density | 27 (4.9) |
| HC_Empfaenge | %Care allowance receivers in all persons in long-term care in 2017 | Care need | 50.86 (6.52) |
| HC_Krankenha | Beds in hospitals per 1000 persons in 2016 | Health | 6.35 (3.89) |
| HC_Pflegebed | Persons in long-term care per 10.000 persons in 2017 | Care need | 428.13 (106.03) |
| HC_Pflegedi | Stuff in care services per 10,000 persons in 2017 | Care need | 47.1 (18.7) |
| HC_Pflegehe | Stuff in nursing homes per 10,000 persons in 2017 | Care need | 97.71 (23.28) |
| HC_Pflegehei | Beds in nursing homes per 100,000 persons in 2017 | Care need | 113.19 (28.86) |
| HC_Stationae | %Persons in inpatient long-term care in all persons in long-term care in 2017 | Care need | 24.36 (5.37) |
| HC_v879 | %Change in beds in hospitals in 2012-2016 | Health | -0.67 (9.1) |
| HC_Vorzeitig | Premature mortality (deaths of persons younger than 65 years) per 1,000 persons in 2017 | Health | 1.72 (.36) |
| inc_14d_274 | Cum. incidence per 100.000 for 14 days in neighbour country at day 274 | Health | 20.84 (44.33) |
| inc_14d_289 | Cum. incidence per 100.000 for 14 days in neighbour country at day 289 | Health | 78.52 (176.94) |
| inc_14d_305 | Cum. incidence per 100.000 for 14 days in neighbour country at day 305 | Health | 193.53 (426.17) |
| inc_14d_320 | Cum. incidence per 100.000 for 14 days in neighbour country at day 320 | Health | 201.51 (400.71) |
| inc_14d_335 | Cum. incidence per 100.000 for 14 days in neighbour country at day 335 | Health | 127.61 (265.31) |
| INC_Bruttov | Gross income per employee in 2016 | SES | 2337.79 (806.78) |
| INC_Bruttow | Gross value added in total in 1,000" per employed person in 2016 | SES | 60.39 (10.78) |
| INC_Haushal | Average household income per person in 2016 | SES | 1591.69 (589.47) |
| INC_Mediane | Median wages of full-time dependently employed persons in 2017 | SES | 2834.29 (882.01) |
| INC_Schuldn | Private debitors per 100 persons in 2017 | SES | 9.69 (2.73) |
| INC_v1161 | GDP in 1,000" per person in 2016 | SES | 35.61 (15.81) |
| INC_v1175 | GDP in 1,000" per employed person in 2016 | SES | 67.05 (11.97) |
| INC_v1176 | %Change in gross value added in 2012-2016 | SES | 13.64 (5.68) |
| INC_v757 | %Change in average household income per person in 2012-2016 | SES | 8.95 (2.96) |
| INC_v775 | %Change of gross income per employee in 2012-2016 | SES | 14.99 (4.46) |
| INC_v792 | Gross income per employee in production industry in 2016 | SES | 3039.73 (1074.39) |
| INC_v793 | %Change in gross income per employee in production industry in 2012-2016 | SES | 11.95 (7.73) |
| INC_v801 | Median wages of full-time dependently employed persons at age 25-55 in 2017 | SES | 2846.08 (926.15) |
| IS_einpen | %Inbound commuters in all employed persons in 2017 | Connectedness | 64.51 (10) |
| IS_endlers~2017 | %Net sum of commuters in all employed persons in 2017 | Connectedness | -10.36 (29.72) |
| IS_mitarbeit~30 | %Change of outbound commuters over a distance of 300km+ in all employed persons in 2007-2017 | Connectedness | 0.02 (0.55) |
| IS_mitarbeit~50 | %Change of outbound commuters over a distance of 50km+ in all employed persons in 2007-2017 | Connectedness | 0.24 (1.41) |
| IS_nAutoba~2018 | Average travel time to the next highway with car in minutes in 2018 | Connectedness | 11.86 (8.37) |
| IS_nFlugha~2018 | Average travel time to the next international airport with car in minutes in 2018 | Connectedness | 49.62 (21.98) |
| IS_Oberzen~2018 | Average travel time to the next large-sized regional center ("Oberzentrum") in 2018 | Connectedness | 22.56 (16.03) |
| IS_Pkw_Di | Cars per 1,000 persons in 2017 | Connectedness | 579.16 (70.98) |
| IS_Strass | Traffic accidents per 100,000 persons in 2017 | Urbanity/density | 491.09 (87.67) |
| IS_v1065 | Average travel time to the next national train station with car in minutes in 2017 | Connectedness | 21.93 (15.38) |
| IS_v1067 | Average travel time to the next medium-sized regional center ("Mittelzentrum") in 2017 | Connectedness | 6.79 (5.55) |
| IS_v1068 | Average distance to the next supermarket (population-weighted in straight-line) in 2017 | Urbanity/density | 1050.97 (542.18) |
| IS_v1069 | %Persons with max. 1km distance to supermarket in 2017 | Urbanity/density | 69.28 (15.82) |
| IS_v1070 | Average distance to the next pharmacy (population-weighted in straight-line) in 2017 | Urbanity/density | 1418.31 (808.55) |
| IS_v1071 | %Persons with max. 1km distance to pharmacy in 2017 | Urbanity/density | 61.76 (18.47) |
| IS_v1072 | Average distance to the next stop/station (population-weighted in straight-line) in 2017 | Connectedness | 531.12 (495.52) |
| IS_v1073 | %Persons with max. 1km distance to stop/station in 2017 | Connectedness | 88.7 (14.05) |
| IS_weg150kmun~r | %Outbound commuters over a distance of 150km+ in all employed persons in 2017 | Connectedness | 4.41 (1.35) |
| IS_weg300kmun~r | %Outbound commuters over a distance of 300km+ in all employed persons in 2017 | Connectedness | 2.4 (.89) |
| IS_weg50kmund~2 | %Outbound commuters over a distance of 50km+ in all employed persons in 2017 | Connectedness | 11.48 (3.48) |
| PO_Alters | %Persons at age 65+ with basic social security benefits in persons aged 65+ in 2017 | SES | 2.62 (1.53) |
| PO_Hausha | %Households with high income (>3,600" per month) in all households in 2016 | SES | 21.17 (5.41) |
| PO_SGBII_ | %Persons with basic social security benefits per 1,000 persons in 2017 | SES | 9 (4.15) |
| PO_v1265 | %Households with average income (1,500" - 3,600" per month) in all households in 2016 | SES | 48.2 (1.58) |
| PO_Versch | Debts of the core households in " per person in 2016 | SES | 1682.7 (1549.75) |
| PRE_Gymnas | %High school students in all pupils in 2017 | SES | 26.36 (6.69) |
| PRE_Roemis | %Roman-catholics in 2011 | Norms and values | 32.24 (24.36) |
| PRE_Schula | %School leavers without any degree in 2017 | SES | 6.56 (2.24) |
| PRE_stimme~2017 | %Valid votes for AfD in all valid votes in 2017 | Norms and values | 13.39 (5.33) |
| PRE_v652 | %Pupils in grade 11 in all pupils in 2017 | SES | 3.81 (1.14) |
| PRE_v675 | %Graduates with secondary education degree in all graduates in 2017 | SES | 16.57 (5.04) |
| PRE_v698 | %Graduates with higher education entrance qualification in all graduates in 2017 | SES | 32.46 (8.86) |
| PRE_v727 | %Employed persons without qualification in all dependently employed persons in 2017 | SES | 11.65 (3.18) |
| PRE_v733 | %Employed persons with academic degree in all dependently employed persons in 2017 | SES | 13.06 (6.2) |
| PRE_v739 | %Employed persons at age 30-35 with academic degree in all dependently employed in 2017 | SES | 2.1 (1.45) |
| PRE_Wahlbe | %Voter turnout (Number of valid votes in the last Bundestag election) of all registered voters in 2017 | Norms and values | 75.08 (3.79) |
| SSE_AnteilErh | %Recreational area in total area in 2017 | Urbanity/density | 2.49 (2.68) |
| SSE_AnteilFre | %Open area (incl. water and agricultural areas) in total area in 2017 | Urbanity/density | 81.22 (12.82) |
| SSE_Anteilnat | %Area in natural state in total area in 2017 | Urbanity/density | 5.1 (3.71) |
| SSE_AnteilWas | %Water area in total area in 2017 | Urbanity/density | 2.4 (2.8) |
| SSE_BetteninF | Beds in tourist facilities per 1,000 persons in 2017 | SES | 41.78 (49.31) |
| SSE_Einwohner | Persons per sqkm in 2017 | Urbanity/density | 490.63 (677.72) |
| SSE_Erholungs | Recreational area in sqm per person in 2017 | Urbanity/density | 70.51 (56.73) |
| SSE_grand_~2016 | Grand mean (unweighted) of PM 10 in 2016 | Urbanity/density | 14.25 (2.2) |
| SSE_Laendlich | %Persons in municipalities with a population density <150 inh/sqkm in 2017 | Urbanity/density | 29.51 (30.14) |
| SSE_Siedlungs | %Persons per sqkm settlement and traffic area in 2017 | Urbanity/density | 1828.86 (1057.55) |
| SSE_Stickstof | Nitrogen surplus per agricultural area in kg/ha in 2016 | Urbanity/density | 68.33 (27.16) |
| SSE_v1130 | %Change in beds in tourist facilities per 1,000 persons in 2012-2017 | SES | -86.78 (6.36) |
| SSE_v833 | Area in natural state in sqm per person in 2017 | Urbanity/density | 2.9 (3.52) |
| SSE_v967 | Persons and employees per sqkm in 2017 | Urbanity/density | 727.79 (1056.47) |
| SSE_v988 | Area size-weighted population in a distance of 100 km in 1,000 persons in 2017 | Urbanity/density | 33367.32 (30681.78) |
| SSE_Waldflaec | Forest area in sqm per person in 2017 | Urbanity/density | 1886.79 (1906.89) |
| SSE_Wasserfla | Water area in sqm per person in 2017 | Urbanity/density | 123.13 (181.27) |
| Std_e00e90end~i | Age standardized rate E00-E99 of Endocrine, nutritional and metabolic diseases per 10,000 persons in 2017 | Health | 64.05 (15.09) |
| Std_i00i99kra~e | Age standardized rate of I00-I99 of diseases of the circulatory system per 10,000 persons in 2017 | Health | 350.95 (70.65) |
| Std_j00j99kra~e | Age standardized rate of J00-J99 of diseases of the respiratory system per 10,000 persons in 2017 | Health | 156.74 (33.15) |
| std_inz_AGS~274 | Age-standardized incidence rate per 100,000 person-years from 16.09. to 30.09.2020 | Health | 26.81 (19.82) |
| std_inz_AGS~289 | Age-standardized incidence rate per 100,000 person-years from 01.10. to 15.10.2020 | Health | 52.45 (36.79) |
| std_inz_AGS~305 | Age-standardized incidence rate per 100.000 person-years from 16.10. to 31.10.2020 | Health | 196.86 (100.08) |
| std_inz_AGS~320 | Age-standardized incidence rate per 100,000 person-years from 01.11. to 15.11.2020 | Health | 294.75 (132.15) |
| std_inz_AGS~335 | Age-standardized incidence rate per 100,000 person-years from 16.11. to 30.11.2020 | Health | 299.44 (144.25) |
| UE_AnteArbei | %Older unemployed (55 years+) in all unemployed persons in 2017 | SES | 22.97 (4.19) |
| UE_AnteBesch | %Older employed persons (55 years+) in all employed persons in 2017 | SES | 20.19 (2.24) |
| UE_AntreArbe | %Young unemployed (under 26 years) in all unemployed persons in 2017 | SES | 9.83 (2.02) |
| UE_AntreBesc | %Young employed persons (under 26 years) in all employed persons in 2017 | SES | 20.86 (2.79) |
| UE_ArbquoteJ | Unemployment rate of young persons (under 26 years) in 2017 | SES | 5.26 (2.71) |
| UE_BeseimHan | Employed persons in crafts sector per 100 employable persons in 2017 | SES | 13.99 (4.89) |
| UE_BesePrima | %Employed persons in primary sector in all dependently employed persons in 2017 | SES | 1.14 (1.27) |
| UE_Dienst | Employed persons in service sector per 100 employable persons in 2017 | SES | 39.24 (14.84) |
| UE_Erwerb | Employed persons per sqm of region in 2016 | Urbanity/density | 327.22 (491.33) |
| UE_Indust | Employed persons in industry per 100 employable persons in 2017 | SES | 18.25 (8.72) |
| UE_Kurzar | %Persons with a short-time job in all employed persons in 2017 | SES | 0.75 (1.44) |
| UE_Langze | %Long-term unemployed persons (unemployed for one year and longer) in 2017 | SES | 32.2 (8.08) |
| UE_QuoBescha | %Older employed persons in all older persons (55 years+) in 2011-2017 | SES | 52.53 (3.6) |
| UE_QuoeBesch | %Young employed persons in all young persons (under 26 years) in 2017 | SES | 49.56 (5.94) |
| UE_v129 | %Change of proportion of young employed persons (under 26 years) in all employed in 2011-2017 | SES | 5.67 (9.24) |
| UE_v166 | %Change of share of older employed persons in all employed persons in 2011-2017 | SES | 38.3 (7.72) |
| UE_v190 | %Persons without any professional qualification in all employed persons in 2017 | SES | 11.71 (3.01) |
| UE_v196 | %Persons with a professional qualification in all employed persons in 2017 | SES | 66.92 (6.63) |
| UE_v202 | %Persons with an academic qualification in all employed persons in 2017 | SES | 11.96 (5.17) |
| UE_v214 | %Change of marginally employed persons in 2012-2017 | SES | 1.27 (7.73) |
| UE_v219 | %Marginally employed persons in all persons 65+ in 2017 | SES | 12.9 (2.77) |
| UE_v220 | %Change of number of marginally employed persons 65+ in 2012-2017 | SES | 34.56 (13.28) |
| UE_v231 | %Marginally employed persons 65+ in all marginally employed persons in 2017 | SES | 15.15 (3.23) |
| UE_v251 | %Employed persons in secondary sector in all dependently employed persons in 2017 | SES | 31.87 (11.46) |
| UE_v305 | %Employed persons in primary sector in all employed persons in 2016 | SES | 2.07 (1.79) |
| UE_v306 | %Change in employed persons in primary sector in all employed persons in 2012-2016 | SES | -6.16 (22.53) |
| UE_v324 | %Change in employed persons in secondary sector in all employed persons in 2012-2016 | SES | 1.13 (5.36) |
| UE_v342 | %Change in employed persons in tertiary sector in all employed persons in 2012-2016 | SES | 3.8 (3.63) |
| UE_v359 | %Employed persons in primary sector in all employed persons in 2016 | SES | 68.93 (11.2) |
| UE_v376 | %Employed persons in finance and housing in all employed in service sector in 2016 | SES | 22.24 (4.54) |
| UE_v381 | %Employed persons in producing professions in all dependently employed persons in 2016 | SES | 30.27 (7.12) |
| UE_v75 | %Change in older age unemployment rate (55 years+) in 2012-2017 | SES | -.34 (15.46) |
| UE_v76 | %Change in younger age unemployment rate (under 26 years) in 2012-2017 | SES | -9.04 (22.95) |
| UE_v97 | %Change of long-term unemployment rate in 2012-2017 | SES | -8.35 (18.69) |

Supplementary Table 2: R^2^ and RMSE scores of boosting models for all periods (Incidence)

|  | 1^st^ Oct - 15^th^ Oct | 16^th^ Oct - 31^th^ Oct | 1^st^ Nov - 15^th^ Nov | 16^th^ Nov - 30^th^ Nov | 1^st^ Dec - 15^th^ Dec |
| --- | --- | --- | --- | --- | --- |
| R^2^ first model (all features) | 0.9994 | 0.9994 | 0.9995 | 0.9993 | 0.9996 |
| RMSE first model (all features) | 0.8771 | 2.4047 | 2.9688 | 3.7350 | 3.7249 |
| R^2^ second model (20 features) | 0.9971 | 0.9963 | 0.9976 | 0.9972 | 0.9983 |
| RMSE second model (20 features) | 1.9621 | 6.0657 | 6.4013 | 7.6349 | 8.0680 |
| R^2^ second model (out-of-sample) | 0.4911 | 0.6369 | 0.7247 | 0.7108 | 0.7428 |
| RMSE second model (out-of-sample) | 26.7168 | 61.2432 | 72.8262 | 79.6925 | 93.5302 |

Supplementary Table 3: R^2^ and RMSE scores of boosting models for all periods (death rates)

|  | 1^st^ Oct - 15^th^ Oct | 16^th^ Oct - 31^th^ Oct | 1^st^ Nov - 15^th^ Nov | 16^th^ Nov - 30^th^ Nov | 1^st^ Dec - 15^th^ Dec |
| --- | --- | --- | --- | --- | --- |
| R^2^ first model (all features) | 0.9990 | 0.9974 | 0.9987 | 0.9987 | 0.9989 |
| RMSE first model (all features) | 0.0279 | 0.1408 | 0.1404 | 0.2841 | 0.3221 |
| R^2^ second model (20 features) | 0.9954 | 0.9918 | 0.9915 | 0.9946 | 0.9949 |
| RMSE second model (20 features) | 0.0598 | 0.2500 | 0.3753 | 0.5752 | 0.6944 |
| R^2^ second model (out-of-sample) | 0.1722 | 0.1814 | 0.2773 | 0.3038 | 0.4213 |
| RMSE second model (out-of-sample) | 0.7927 | 2.6000 | 3.6652 | 6.0477 | 6.9774 |

Supplementary Figure 1: Period 1 - Regional distribution of age-standardized COVID-19 incidence and death rates

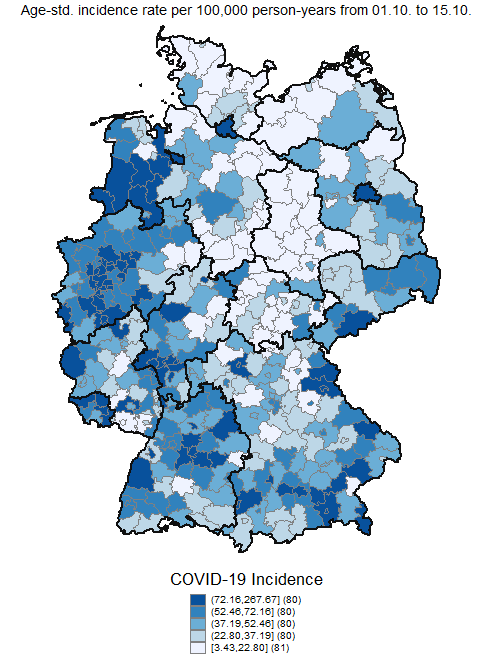

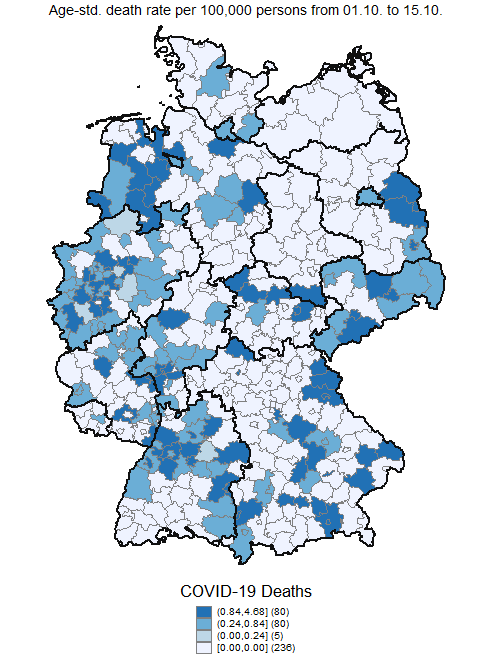

Supplementary Figure 2: Period 2 - Regional distribution of age-standardized COVID-19 incidence and death rates

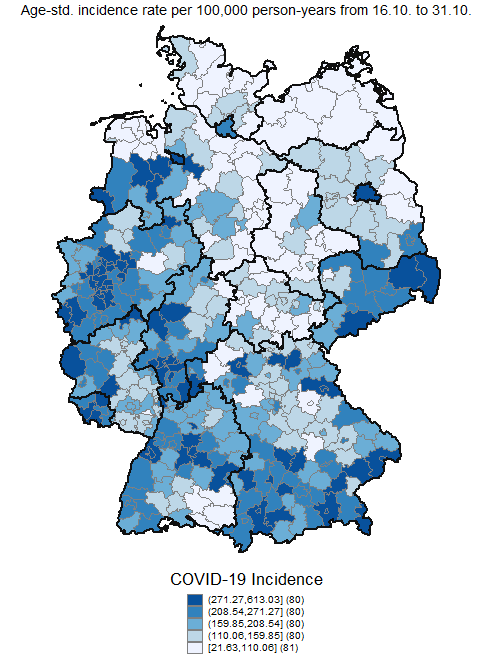

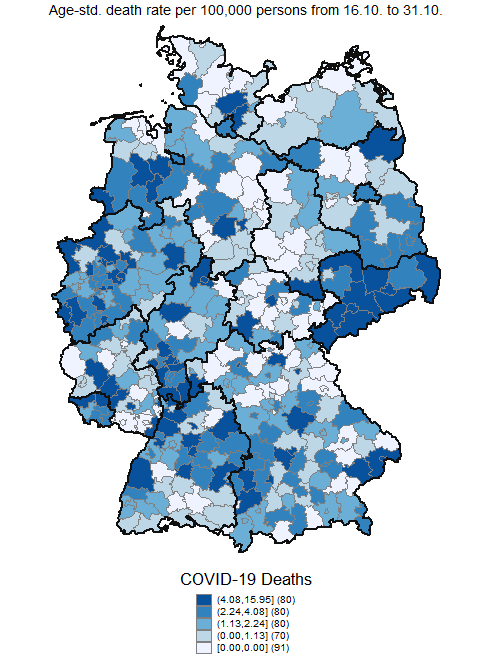

Supplementary Figure 3: Period 3 - Regional distribution of age-standardized COVID-19 incidence and death rates

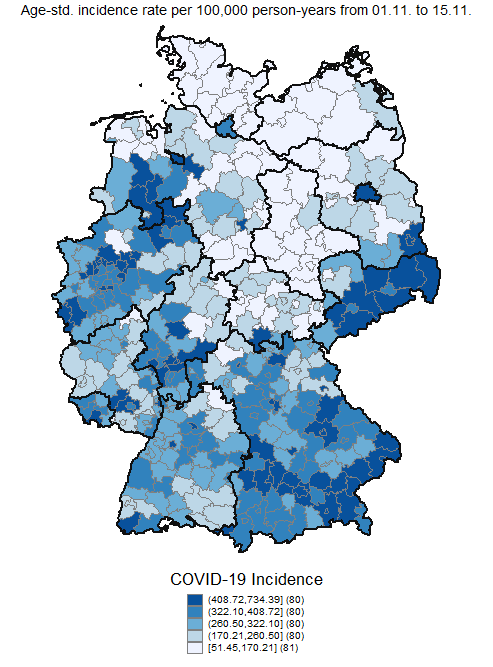

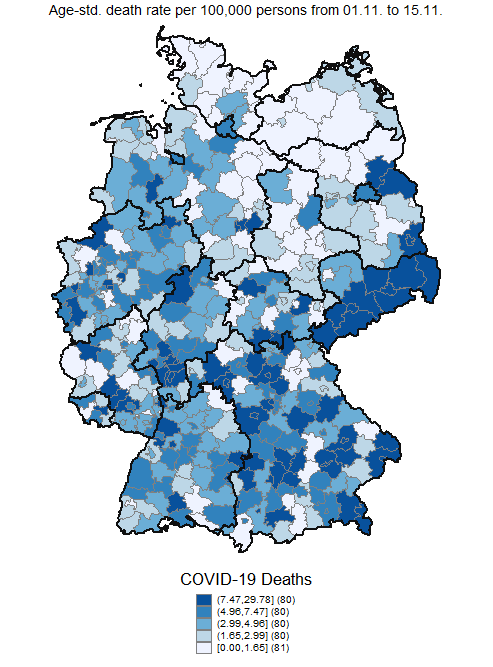

Supplementary Figure 4: Period 4 - Regional distribution of age-standardized COVID-19 incidence and death rates

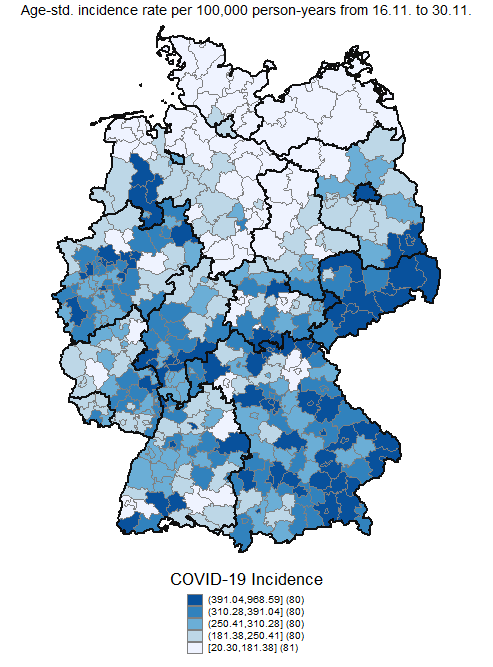

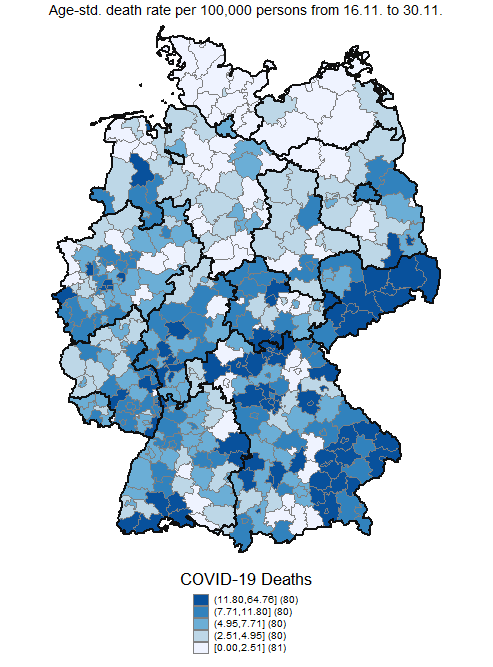

Supplementary Figure 5: Period 5 - Regional distribution of age-standardized COVID-19 incidence and death rates

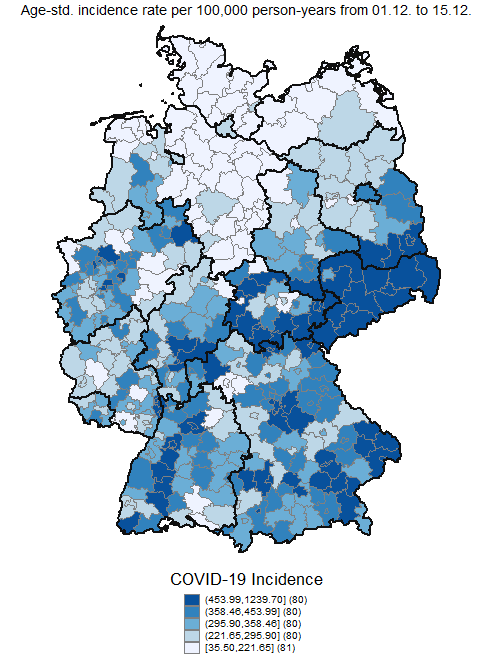

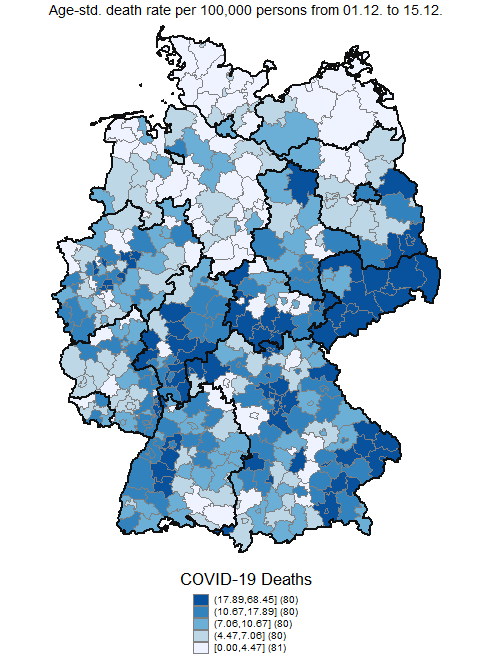

Supplementary Figure 6: Period 1 - SHAP summary plots of the first twenty features (a) age-standardized incidence, (b) age-standardized death rates

(b)

(a)

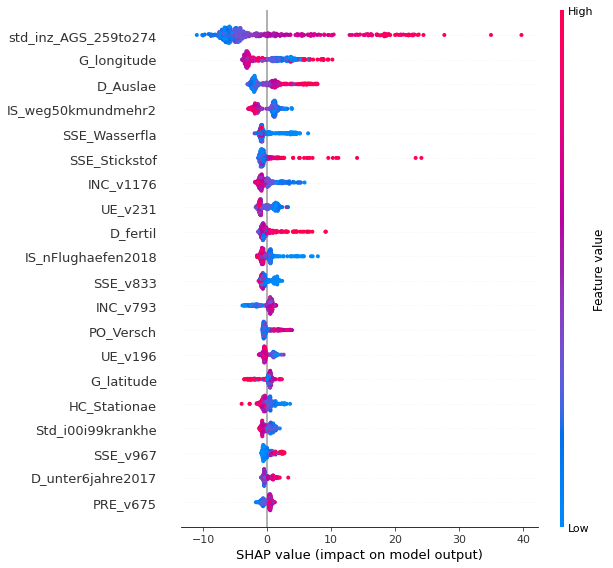

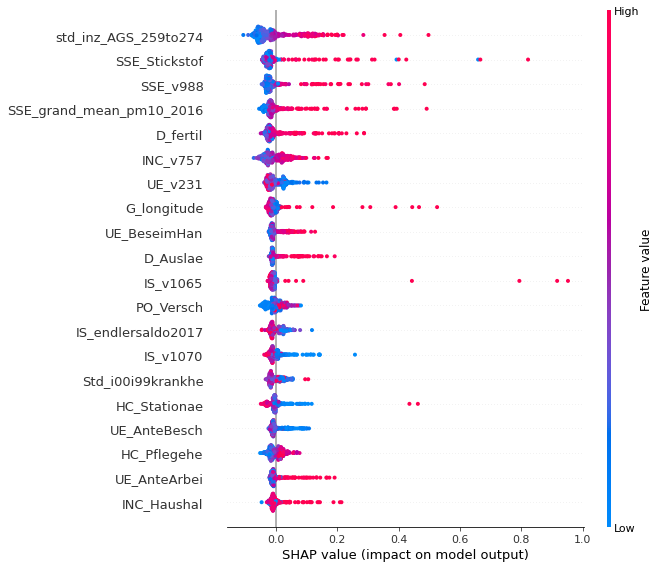

The summary plot combines feature importance with feature effects. Each point on the summary plot is a Shapley value for a feature and a county. The position on the y-axis is determined by the feature and on the x-axis by the Shapley value. The color represents the value of the feature from low to high. Overlapping points are jittered in y-axis direction, to get a sense of the distribution of the Shapley values per feature. The features are ordered according to their importance. E.g. Low values of the age-standardized incidence in the previous period (std_inz_AGS_259to274) are correlated with low values in the age-standardized incidence of the current period (a). High values of longitude (G_longitude) are correlated with low values of the age-standardized incidence of the current period (a). High levels of “Nitrogen surplus per agricultural area in kg/ha in 2016” (SSE_Stickstof) are correlated with high age-standardized death rates in the current period (b). For the labels of the features see Supplemental Table 1.

Supplementary Figure 7: Period 2 - SHAP summary plot of the first twenty features (a) age-standardized incidence, (b) age-standardized death rates

(b)

(a)

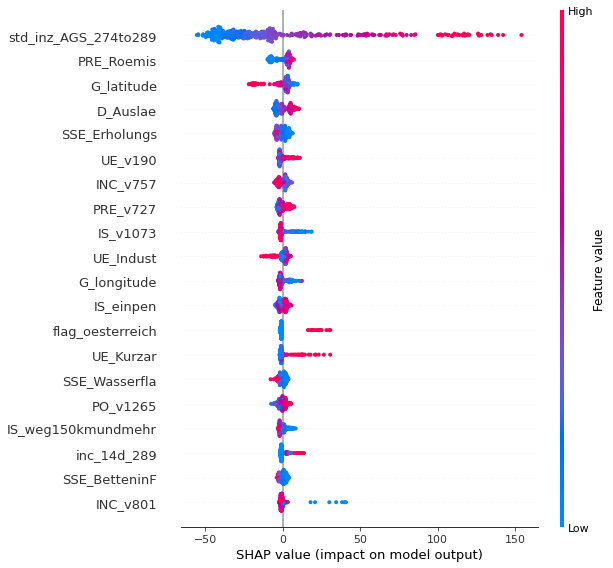

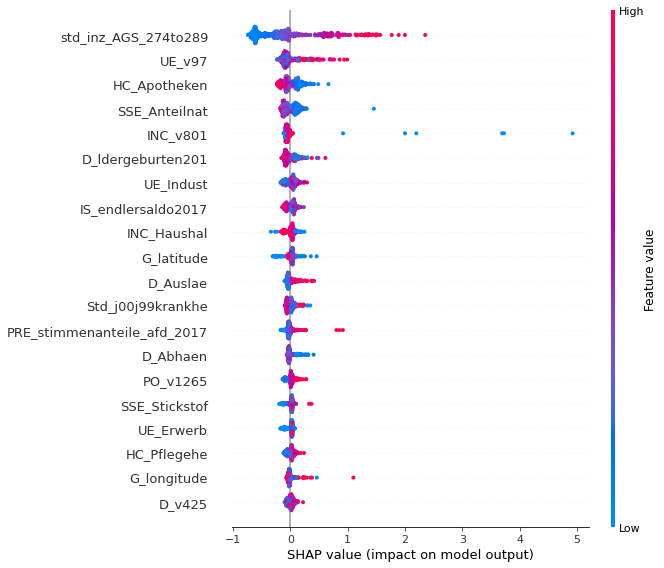

The summary plot combines feature importance with feature effects. Each point on the summary plot is a Shapley value for a feature and a county. The position on the y-axis is determined by the feature and on the x-axis by the Shapley value. The color represents the value of the feature from low to high. Overlapping points are jittered in y-axis direction, to get a sense of the distribution of the Shapley values per feature. The features are ordered according to their importance. E.g. Low values of the age-standardized incidence in the previous period (std_inz_AGS_274to289) are correlated with low values in the age-standardized incidence (a) and death rate(b) of the current period. A large proportion of people with Roman-Catholic denomination (Pre_Roemis) is correlated with low values of the age-standardized incidence of the current period (a). Large “%Changes of the long-term unemployment rate in 2012-2017” (UE_v97) are correlated with high values of age-standardized death rates in the current period (b). For the labels of the features see Supplemental Table 1.

Supplementary Figure 8: Period 3 - SHAP summary plot of the first twenty features (a) age-standardized incidence, (b) age-standardized death rates

(a)

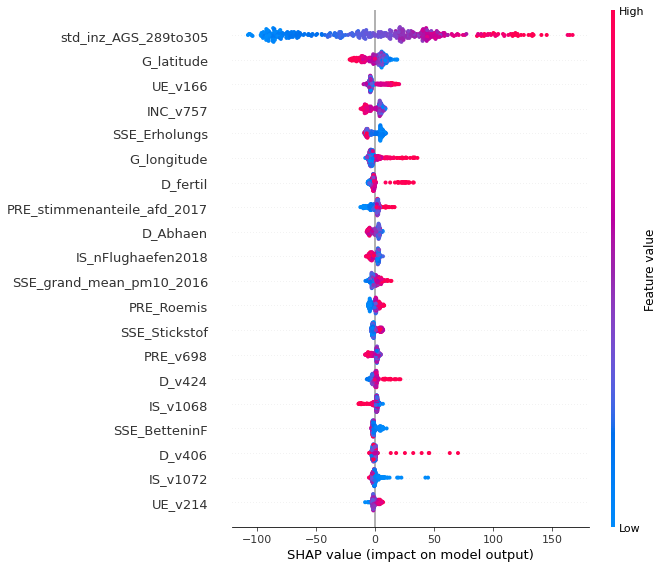

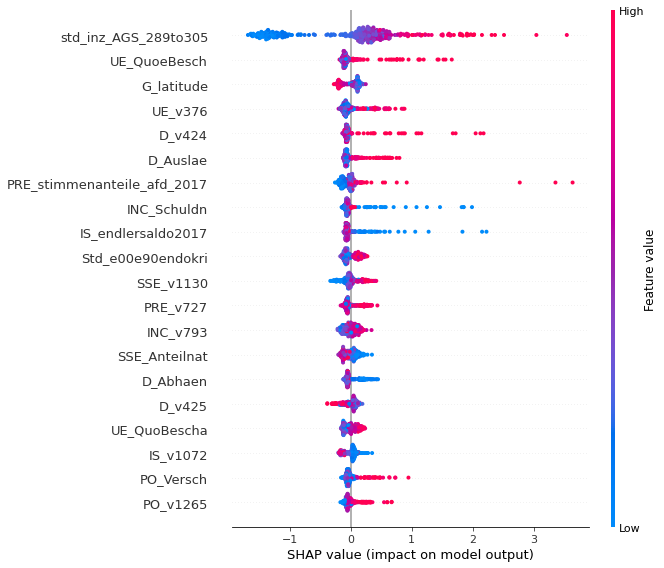

(b)

The summary plot combines feature importance with feature effects. Each point on the summary plot is a Shapley value for a feature and a county. The position on the y-axis is determined by the feature and on the x-axis by the Shapley value. The color represents the value of the feature from low to high. Overlapping points are jittered in y-axis direction, to get a sense of the distribution of the Shapley values per feature. The features are ordered according to their importance. E.g. Low values of the age-standardized incidence in the previous period (std_inz_AGS_289to305) are correlated with low values in the age-standardized incidence (a) and death rate(b) of the current period. A large “%Change of share of older employed persons in all employed persons in 2011-2017” (UE_v166) is correlated with low values of the age-standardized incidence of the current period (a). A large “%Young employed persons in all young persons (under 26 years) in 2017” (UE_QuoeBesch) is correlated with high values of age-standardized death rates in the current period (b). For the labels of the features see Supplemental Table 1.

Supplementary Figure 9: Period 4 - SHAP summary plot of the first twenty features (a) age-standardized incidence, (b) age-standardized death rates

(b)

(a)

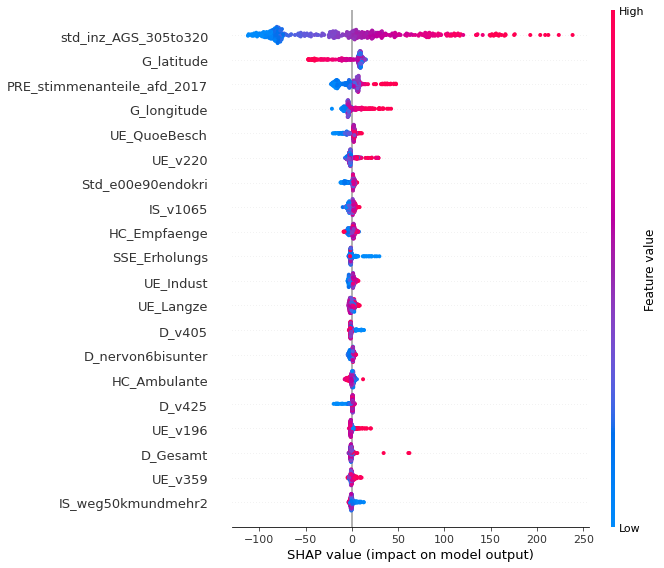

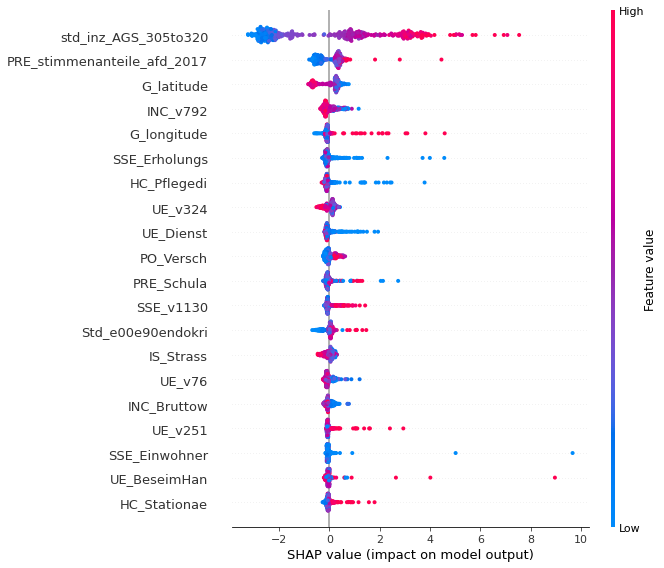

The summary plot combines feature importance with feature effects. Each point on the summary plot is a Shapley value for a feature and a county. The position on the y-axis is determined by the feature and on the x-axis by the Shapley value. The color represents the value of the feature from low to high. Overlapping points are jittered in y-axis direction, to get a sense of the distribution of the Shapley values per feature. The features are ordered according to their importance. E.g. Low values of the age-standardized incidence in the previous period (std_inz_AGS_305to320) are correlated with low values in the age-standardized incidence (a) and death rate(b) of the current period. A large “%Valid votes for AfD in all valid votes in 2017” (PRE_stimmenanteile_afd_2017) is correlated with high values of the age-standardized incidence (a) and death rate (b) of the current period (a). For the labels of the features see Supplemental Table 1.

Supplementary Figure 10: Period 5- SHAP summary plot of the first twenty features (a) age-standardized incidence, (b) age-standardized death rates

(b)

(a)

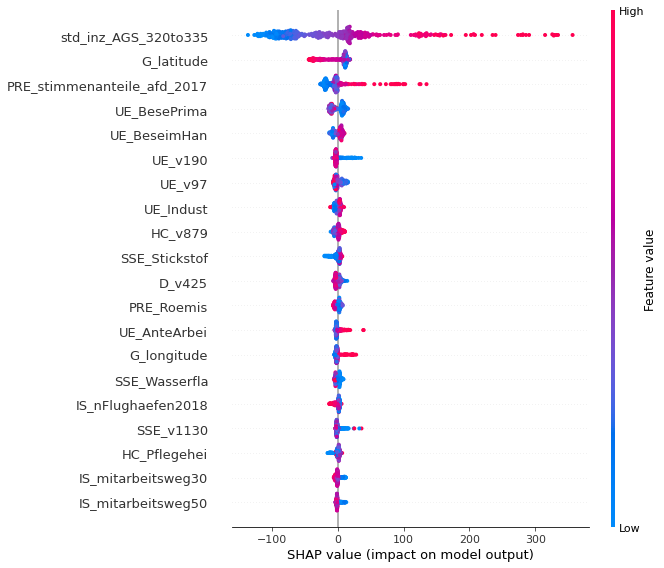

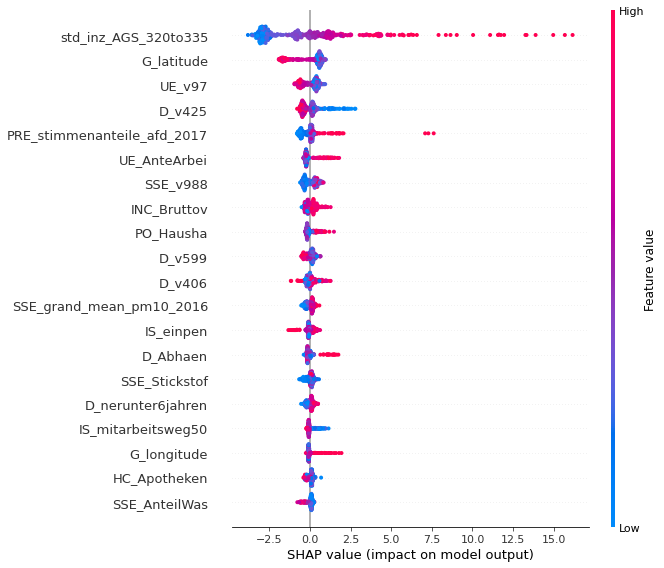

The summary plot combines feature importance with feature effects. Each point on the summary plot is a Shapley value for a feature and a county. The position on the y-axis is determined by the feature and on the x-axis by the Shapley value. The color represents the value of the feature from low to high. Overlapping points are jittered in y-axis direction, to get a sense of the distribution of the Shapley values per feature. The features are ordered according to their importance. E.g. Low values of the age-standardized incidence in the previous period (std_inz_AGS_320to335) are correlated with low values in the age-standardized incidence (a) and death rate(b) of the current period. A large “%Valid votes for AfD in all valid votes in 2017” (PRE_stimmenanteile_afd_2017) is correlated with high values of the age-standardized incidence (a) and death rate (b) of the current period (a). For the labels of the features see Supplemental Table 1.
